# Performance and Practicality of 16S Nanopore Sequencing for Routine Bacterial Identification in Clinical Samples

**DOI:** 10.64898/2025.12.04.25341610

**Authors:** Authors: A.U. Geers, C. Bütikofer, M. A. Terrazos Miani, S. Droz, A. Zihler Berner, I. Lendenmann, P.M. Keller, F. Suter-Riniker, S. Neuenschwander, C. Hirzel, C. Casanova, A. Ramette

## Abstract

**Background:** The accurate and timely identification of bacterial pathogens in low-diversity samples is critical for clinical diagnostics, yet traditional culture-based methods often fail due to prior antibiotic exposure or fastidious growth requirements. While 16S rRNA gene sequencing provides a culture-independent alternative, traditional Sanger sequencing cannot resolve polymicrobial infections, and short-read sequencing platforms are often limited by long turnaround time and high associated costs. Oxford Nanopore Technologies (ONT) offers a promising solution through rapid turnaround times and lower costs; however, its clinical adoption is hindered by a lack of standardized, validated protocols and large-scale comparative data.

**Methods:** We developed a rapid, cost-effective 16S rRNA diagnostic workflow using ONT and benchmarked its performance against an Illumina Next-Generation Sequencing (NGS) workflow. The pipeline utilizes an *in vitro* diagnostic (IVD) certified amplification protocol targeting the V3-V4 region with DNA-free reagents to minimize contamination in low-biomass specimens. We first verified the detection limits using dilution series of pure and mixed bacterial cultures. Subsequently, the workflow was prospectively applied to 101 clinical samples, with results evaluated retrospectively against Illumina data and subjected to rigorous clinical review to determine infection plausibility.

**Results:** Comparative analysis revealed high concordance between NGS platforms across the clinical cohort, achieving a weighted species overlap of 93.5 ± 7.6% and a Cohen’s *kappa* of 0.81 ± 0.04 upon clinical review. While Illumina demonstrated slightly higher sensitivity at the lowest microbial concentrations in dilution series experiments, ONT generated sequences with comparable average accuracy (99.9 ± 0.39% for ONT, 99.8 ± 0.42% for Illumina). Crucially, the ONT workflow was significantly more efficient, requiring approximately 50 hours less total processing time and proving more cost-effective for small-batch, on-demand diagnostics than the Illumina workflow.

**Conclusions:** This study offers strong supporting evidence for the integration of 16S nanopore sequencing into routine infectious disease diagnostics. Our findings demonstrate that 16S nanopore sequencing is a feasible, time-efficient, and high-resolution alternative to established NGS methods, particularly suited for rapid, decentralized clinical implementation for diagnostic sequencing of low-diversity samples.

## Introduction

The identification of bacterial pathogens is a critical step in clinical diagnostics, guiding effective treatment and improving patient outcomes. Traditionally, this process has relied on culture-based methods, which can be time-consuming and may fail, due to factors such as prior antibiotic exposure (1) or fastidious growing organisms (2). In recent years, culture-independent approaches, particularly sequencing of the 16S rRNA gene, have become more widely adopted in clinical settings due to their ability to provide rapid taxonomic identification directly from patient samples (3–6). This approach is especially advantageous for complex and urgent cases where timely results are crucial or for samples that yield inconclusive or negative results with conventional culture. Particularly challenging to diagnose are samples with low-bacterial load, such as specimens from sterile body sites, since reagent contamination can lead to significant over estimation of potential pathogens (7). Thus, using DNA-free reagents is crucial to limit the detection of false positives. Several sequencing technologies are available for 16S rRNA gene amplicon sequencing. While traditional Sanger sequencing remains widely used due to its low cost and speed, its inability to resolve complex mixtures of bacteria limits its effectiveness for polymicrobial infections (3, 4). Next-generation sequencing (NGS) platforms, such as those from Illumina, offer high precision, high throughput and the ability to resolve highly complex microbiomes and polymicrobial infections. However, the short read lengths produced by Illumina can sometimes limit the ability to achieve species-level taxonomic resolution, particularly for closely related bacteria (2). Additionally, their high hardware costs and the requirement for large sample batches to be cost-effective are barriers to implementation in a rapid, on-demand clinical diagnostic workflow.

Third-generation sequencing technologies, most notably Oxford Nanopore Technologies (ONT), present a promising alternative. Nanopore sequencing offers advantages that are particularly well-suited for clinical microbiology, including rapid sample-to-result turnaround times, low initial hardware expenditure, and the ability to process single or small batches of samples efficiently (8, 9). Its portability allows for decentralized testing, further reducing time to diagnosis (10, 11). While ONT sequencing has been associated with a higher rate of sequencing errors compared to Illumina, its capacity to generate long reads spanning the full-length 16S rRNA gene can offer superior taxonomic resolution (8). Despite its potential, the clinical adoption of 16S nanopore sequencing is hindered by the lack of standardized and clinically validated protocols. A comprehensive and direct comparison to established NGS methods like Illumina, especially using a large cohort of clinical samples, has also been lacking. Our study addresses these critical gaps by presenting a clinically tested and used pipeline for 16S nanopore sequencing in routine infectious disease diagnostics. We implemented a cost-and time-effective pipeline utilizing an *in vitro* diagnostic (IVD) certified amplification protocol that targets the V3-V4 region of the 16S rRNA gene using DNA-free reagents. The amplification protocol has been previously evaluated on over 800 clinical specimens in conjunction with Sanger sequencing and compared to culturing, yielding an added benefit for the patient in that 36.5% of the PCR-positive results were negative by culture (7). This IVDR-certified kit was used for all steps in the study. We first verified the detection limits using dilution series of pure and mixed bacterial cultures. Subsequently, we prospectively applied the ONT NGS workflow to 101 low-biomass clinical specimens from normally sterile body sites, benchmarking the results against an Illumina NGS workflow, culture findings, and comprehensive clinical reviews to determine infection plausibility.

## Methods

### Clinical samples and pure bacterial cultures

Clinical samples originated from the Insel Gruppe, the largest Swiss hospital group, and were analyzed at the Institute for Infectious Diseases (IFIK) in Bern. The diagnostic division of IFIK (clinical microbiology) is accredited under EN ISO/IEC 17025 to conduct routine bacterial diagnostics on clinical samples. Clinical samples from normally sterile body sites were routinely analyzed with eubacterial 16S PCR and ONT sequencing as described below. Samples for which the patient had provided general consent and which exhibited a DNA concentration of >1 ng/µl DNA after the 16S PCR were subjected to retrospective sequencing with Illumina as described below. The Ethics Committee of the Canton of Bern (BASEC-Nr 2024-00640) approved this study. The independence of observations was assumed for all samples, as the anonymization process precluded the identification of repeated measures from the same patient. As part of the routine diagnostic procedure, culturing from the specimens was systematically attempted under both aerobic and anaerobic conditions for all clinical specimens. For dilution series experiments, *S. aureus* (ATCC 25923, five 16S rRNA operon copies) and *E. coli* (ATCC 25922, seven 16S rRNA operon copies) were grown in 5 ml Luria Broth (LB) liquid culture for 2 h, and subsequently diluted to 10^-5^, 10^-6^, and 10^-7^ in LB. Additionally, undiluted *E. coli* and *S. aureus* liquid cultures were mixed in a ratio of 2:1. The resulting mixture was diluted to 10^-5^, 10^-6^, and 10^-7^ in LB (**Figure 1**). One ml of each of the diluted liquid cultures was used for further DNA extraction and sequencing (see below), and the liquid cultures were plated on MAC (MacConkey II Agar, Becton Dickinson) and CNA (Columbia CNA Agar, Becton Dickinson) plates to determine bacterial counts (colony forming units; CFU).

**Figure 1.**
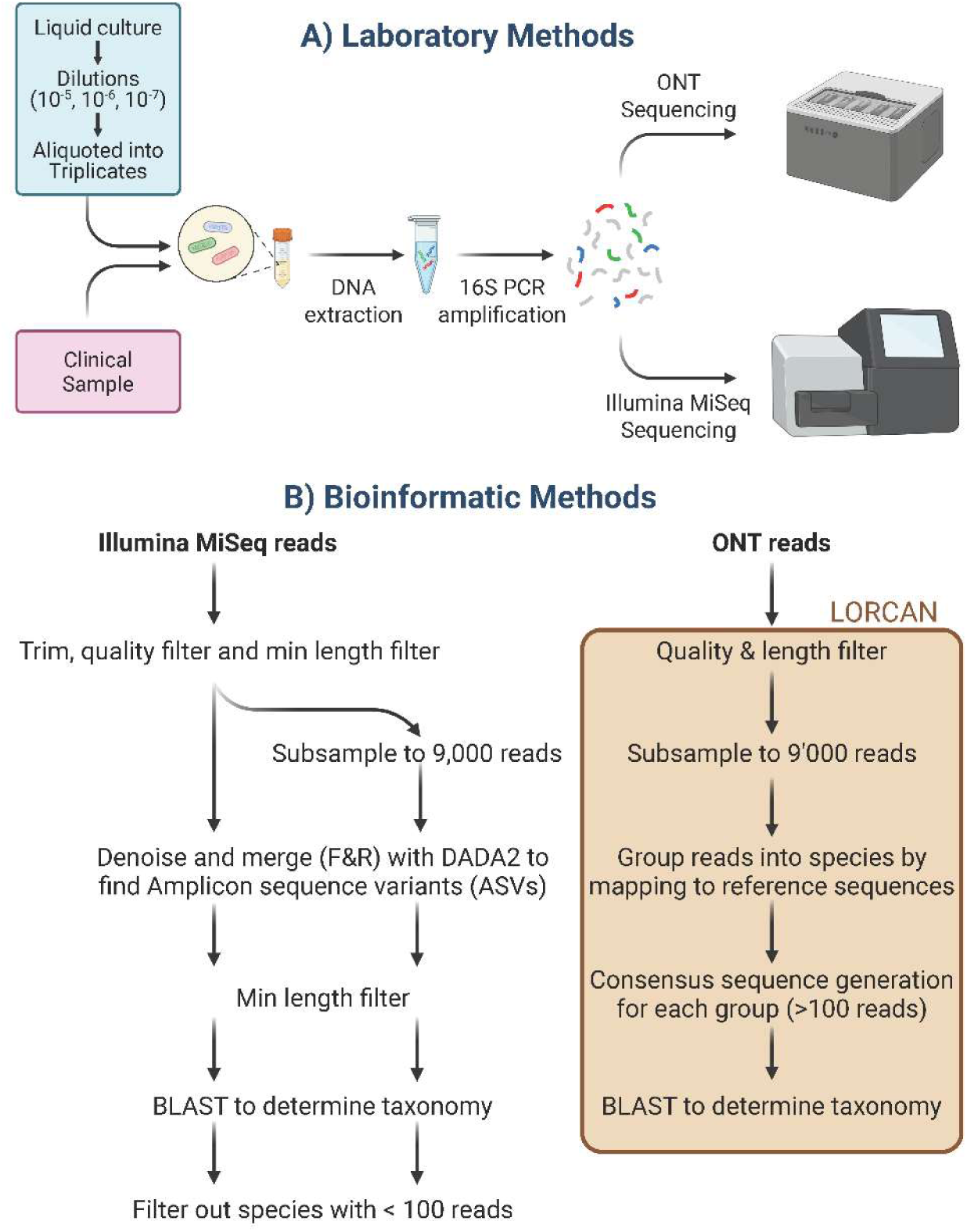
Overview of the laboratory and bioinformatic workflows. **A**) Investigated were dilutions series of pure liquid cultures of *E. coli* and *S. aureus*, as well as over 100 clinical samples from routine diagnostics. The DNA was extracted and the 16S rRNA amplified with the Molzym Micro-Dx kit. The resulting amplicons were sequenced with both ONT and Illumina MiSeq. **B**) All raw sequencing data was quality and length filtered. Illumina reads were additionally subsampled to 9,000 reads before further processing. The DADA2 tool was used to generate amplicon sequence variants (ASVs) from the Illumina reads and the LORCAN pipeline was used for the ONT reads. The taxonomy of the consensus sequences was determined using BLAST. For further analysis only species associated with more than 100 reads were kept. The Figure was created with BioRender https://BioRender.com.

### DNA extraction and PCR amplification

The commercial Micro-Dx kit (Molzym, Bremen, Germany) was used for automated microbial DNA enrichment, host depletion, microbial DNA extraction and real-time 16S PCR (**Figure 1**). The kit was selected on the basis of its prior implementation in our diagnostic laboratory workflow, its IVD-R certification, its DNA free reagents, and its demonstrated high concordance with culture results (7). The manufacturer’s protocol was followed and DNA extractions were done using the manufacturer’s SelectNA plus robotic system. PCR mixtures (28 µl) were assembled with 10 µl master mix containing 7.5 µl water, 2.5 µl DNA staining solution and 0.8 µl MolTaq 16S (DNA polymerase; Molzym). Real-time 16S PCR was performed in a LightCycler z480 (Roche) with the following parameters: 95°C for 1 min; 40 cycles of 95°C for 5 s, 55°C for 10 s, 72°C for 25 s; and 40°C for 10 s. PCR products were purified with AMPure XP beads (Beckman coulter, Brea, California) following the manufacturer’s protocol with minor modifications: An additional 3 s centrifugation step was added after the washing step, and the purified DNA was eluted in 80 µl of Tris-HCl (0.01 M, pH 8.0). Amplicon fragment sizes were assessed using the TapeStation D1000 assay (Agilent, Santa Clara, CA, USA), and DNA concentrations were determined with the Qubit double-stranded DNA (dsDNA) broad-range (BR) assay (Thermo Fisher Scientific, Reinach, Switzerland). Clinical samples that exhibited an identifiable peak in the melting curve at the correct sequence length were subsequently subjected to ONT routine sequencing. All samples from the dilution series were sequenced, irrespective of the melting curve or DNA concentration.

To assess potential contaminations, we used a total of 10 negative controls (i.e. sterile double-distilled water). At the DNA extraction step, five negative controls were set up; and at the eubacterial PCR step further five controls were set up. We further added four negative controls during the library prep Illumina steps. All negative controls were handled together with the samples from their point of set up until the final sequencing step. We were only able to successfully identify species in three negative controls (one extraction negative, two eubacterial PCR negative controls). The species identified in the extraction negative or PCR negative controls could not be identified in samples from the same extraction and PCR batch. The exception was a species of *Actinomycetales* bacterium in one extraction negative control, which could be found with less than 5% relative abundance in one clinical sample (**Supplementary Figure 1**). Taxa found in negative controls were not removed from samples for statistical analysis and clinical reporting. Clinical reports included the results of the negative controls.

### Library preparation and sequencing

Library preparation for ONT sequencing was performed with the kit SQK NBD114.24 (ONT, Oxford, UK) using the supplementary reagents NEBNext Ultra II End repair/dA tailing Module (New England Biolabs [NEB], ON, CA), NEB Blunt/TA Ligase Master Mix (M0367; NEB) and NEBNext Quick Ligation Module (NEB). The manufacturer’s protocol (NBA_9168_v114_revO_15Sep2022) was modified as follows: Diluted DNA Control Sample (DCS) was not used for End Prep (EP) step, DCS volume was substituted with nuclease free water (NFW). No clean-up and no normalization were performed after EP. Therefore, 3 µl of End Prep product with 4.5 µl NFW was used for barcoding step instead of 7.5 normalized EP product. Ethanol (80%) was used for the post-barcoding washes instead of Short Fragment Buffer (SFB). SFB was used for clean-up step after adapter ligation. A typical library prepared under routine conditions consisted of the pooling of PCR amplicons from 1 to 20 clinical samples, two positive controls (*Streptococcus agalactiae* amplicons), one extraction negative and one PCR negative samples. ONT libraries were sequenced on GridION X5 sequencers with the standard flowcells (FLO-MIN114, R10.4.1) with real-time basecalling under high accuracy mode (HAC, basecaller dorado versions 7.1.4-7.6.7), for up to 1 h or until the positive controls reached at least 10,000 reads.

Library preparation for Illumina sequencing was performed with the KAPA Unique Dual-Indexed Adapter Kit KR1736 – v2.19 and the KAPA HyperPrep Kit KR0961 – v7.19. The protocol was followed according to the manufacturer’s instructions, with the exception of using CleanNGS DNA Beads instead of the KAPA cleanup beads, and two additional clean-up steps with beads after amplification. Each library consisted of 68 to 76 pooled clinical samples and negative controls. The library was sequenced on an Illumina MiSeq sequencer with v2 reagents in 2×250 bp paired-end mode. Both ONT and Illumina sequencing were performed at the Institute for Infectious Diseases, University Bern.

### Bioinformatic analyses

ONT reads were demultiplexed and analyzed with the LORCAN pipeline (12). In brief, the adapters are trimmed, filtered by length (± 100 bp around modal sequence length; i.e. 320-520 bp), and subsampled to a maximum of 9,000 reads (**Figure 1**). Then the reads are grouped into species by mapping to a custom nonredundant reference database (see below). Only species groups with at least 100 reads are further processed. For each species group, the reads are remapped to the reference sequence with the highest number of mapped reads, and the consensus sequence is generated using a 50% majority rule consensus. In a last step, the consensus sequences are mapped again to the custom database using BLAST (v2.12.0) to identify the closest taxonomic neighbors (**Figure 1**). Detailed instructions for database creation can be found online: https://github.com/aramette/LORCAN/. In brief the reference database, LeBiBi SSU-rDNA-mk45 (13) was trimmed to the region of interest, dereplicated (mothur; version v.1.48.0 (14)), and used to make a custom database based on the region of interest.

The basecalled and demultiplexed Illumina reads were trimmed, quality filtered (fastp v0.23.2, enabled base correction in overlapped regions, Phred quality ≥ 20), end trimmed (24 bp) and filtered for minimum length of 240 bp (DADA2, v1.34 (15)). Subsequently the reads were denoised, merged and amplicon sequencing variants (ASVs) generated using the established DADA2 (v1.3.4) pipeline (15). The resulting ASVs were again filtered for a minimum length of 390 bp and maximum of 450 bp, and their taxonomy was determined by mapping to the reference database using BLAST (v2.12.0). In a last step read counts were combined at species level and all species with less than 100 reads were filtered out (**Figure 1**). Read counts were processed with R version (4.4.1), using the packages tidyverse (v2.2.0), data.table (v.17.2), RColorBrewer (v1.1-3), ggvenn (v0.1.10), ggpubr (v0.6.0), ggtext (v0.1.2), svglite (v2.21), to generate the figures and perform statistical analyses.

### Clinical review and plausibility assessment

The final sequencing results obtained from both ONT and Illumina, consisting of lists of species that were detected with more than 100 reads in a sample, were evaluated by an experienced infectiologist from the Inselspital, Bern. Similarly to previous studies (16), the expert was tasked with providing an overall assessment of the plausibility of the reported species for the individual samples, as opposed to evaluating each species individually. They were also asked to indicate whether they deemed the sequencing results of each method to be plausible in light of the patient’s medical history, including underlying primary diseases, surgical interventions, complications, microbiological and culturing results from this and other specimens.

### Data availability

Raw sequencing data is available under the BioProject accession number: PRJEB102560. Supplementary tables cited in the text can also be accessed at https://zenodo.org/records/17811713

## Results

### Verification experiments

We compared the detection capabilities of ONT and Illumina sequencing on pure liquid cultures of *E. coli* and *S. aureus*, and on a mixture of the two at a ratio of 2:1, each diluted to 10^−5^, 10^−6^, and 10^−7^ in triplicates, corresponding to average CFU counts of 3,040, 310, 30 for *E. coli*, and 2,590, 80, 0 for *S. aureus*, respectively. These concentrations were selected to be close to the limit of detection observed during the initial testing of the amplification kit (data not shown). In the mixture, there were an average of 34,270, 8,330, 2,110 *E. coli* CFU and 14,600, 6,150, 910 *S. aureus* CFUs in the respective dilutions. All samples were sequenced with both technologies. Additionally, we analyzed a subsample of 9,000 Illumina reads per sample, which represents the maximum number of reads considered in our routine ONT analyses, to assess the impact of sequencing depth on the results. On average we obtained 1,040 raw reads with ONT sequencing and 186,040 raw reads with Illumina sequencing for our dilution series (**Supplementary Table 1**). In the bioinformatic analysis, sequences identified as *Escherichia* sp. and *Shigella* sp. were grouped together due to the high genetic similarity of their 16S rRNA gene and the resulting inability of V3-V4 16S sequencing to distinguish them (17). Similarly, we made the decision to regroup sequences for *Corynebacterium* spp. together. At the highest concentrations, both sequencing methods, with either the full or subsampled read sets, successfully detected the correct species, confirming their ability to identify species present at high concentrations (**Figure 2**). However, at lower concentrations (<310 CFU/ml), neither method could detect *E. coli*, suggesting that the true concentration of this species fell below the limit of detection for both sequencing platforms under these experimental conditions. At these lower bacterial loads, the DNA concentrations after PCR were also below 1 ng/µl and no discernible peak in the melting curve was visible (data not shown). For *S. aureus*, detection failed to be reliable at concentrations below 80 CFU/ml; only Illumina sequencing (including the subsampled data) was able to identify the species at the 10^-6^ dilution in 2 out of 3 replicates, indicating that this method achieved slightly higher analytical sensitivity in detecting low microbial loads. This again corresponded to the measured DNA concentration and the melting curve. In the polymicrobial sample, both methods detected both species even at the lowest concentrations. The observed ratio between the two species was in the same order of magnitude as for the CFU counts. When using ONT sequencing on dilutions 10^−5^, 10^−6^, and 10^−7^, we observed ratios between *E. coli* and *S. aureus* of 8, 3 and 7 in read numbers, respectively. With Illumina sequencing these ratios were 4, 2 and 5, respectively (**Figure 2**). Subsampled Illumina reads had no effect on these ratios. Ratios determined by CFU counts were 2.3, 1.3 and 2.3, respectively. This indicates that although both methods can accurately identify species within a mixed community, minor differences in the observed abundances may occur. Important to consider here is that Gram-negative organisms, such as *E. coli*, are generally easier to lyse and extract than their Gram-positive counterparts, and thus their abundance may be overestimated by sequencing, which could contribute to the skewed observed ratios. A notable finding was the more frequent detection of false positives using Illumina sequencing with the DADA2 pipeline. These false positives included *Paracoccus hibiscisoli*, *Rhizobium daejeonense*, *Corynebacterium* sp., *Streptococcus parasanguinis*, and *Cutibacterium acnes*, among others. False positive detections occurred in both the full or subsampled datasets, indicating that the issue was not related to sequencing depth. We further investigated the ONT raw reads for the presence of these contaminants, and found them to be present as well, albeit below the threshold required (100 reads) for generating consensus sequences (**Supplementary Table 2**). To further evaluate the analytical specificity of the ONT workflow, we also tested a threshold of a minimum of 50 reads for a species to be identified as present in a sample. This resulted in the detection of the contaminants *P. hibiscisoli*, *Corynebacterium* sp., *Erwinia psidii*, *Actinomyces oris*, and *C. acnes* by the ONT workflow, of which the majority were also detected by Illumina at both thresholds. Additionally, the lowering of the threshold resulted in the detection of the expected *S. aureus* at 80 CFU/ml input concentration. However, even at that threshold, Illumina detected contaminations in three times as many samples as ONT. The 100 reads threshold had been previously evaluated on amplicons from clinical cultures (12) and was chosen because it represented a good tradeoff between analytical sensitivity and specificity, which is also evidenced in the present study.

**Figure 2.**
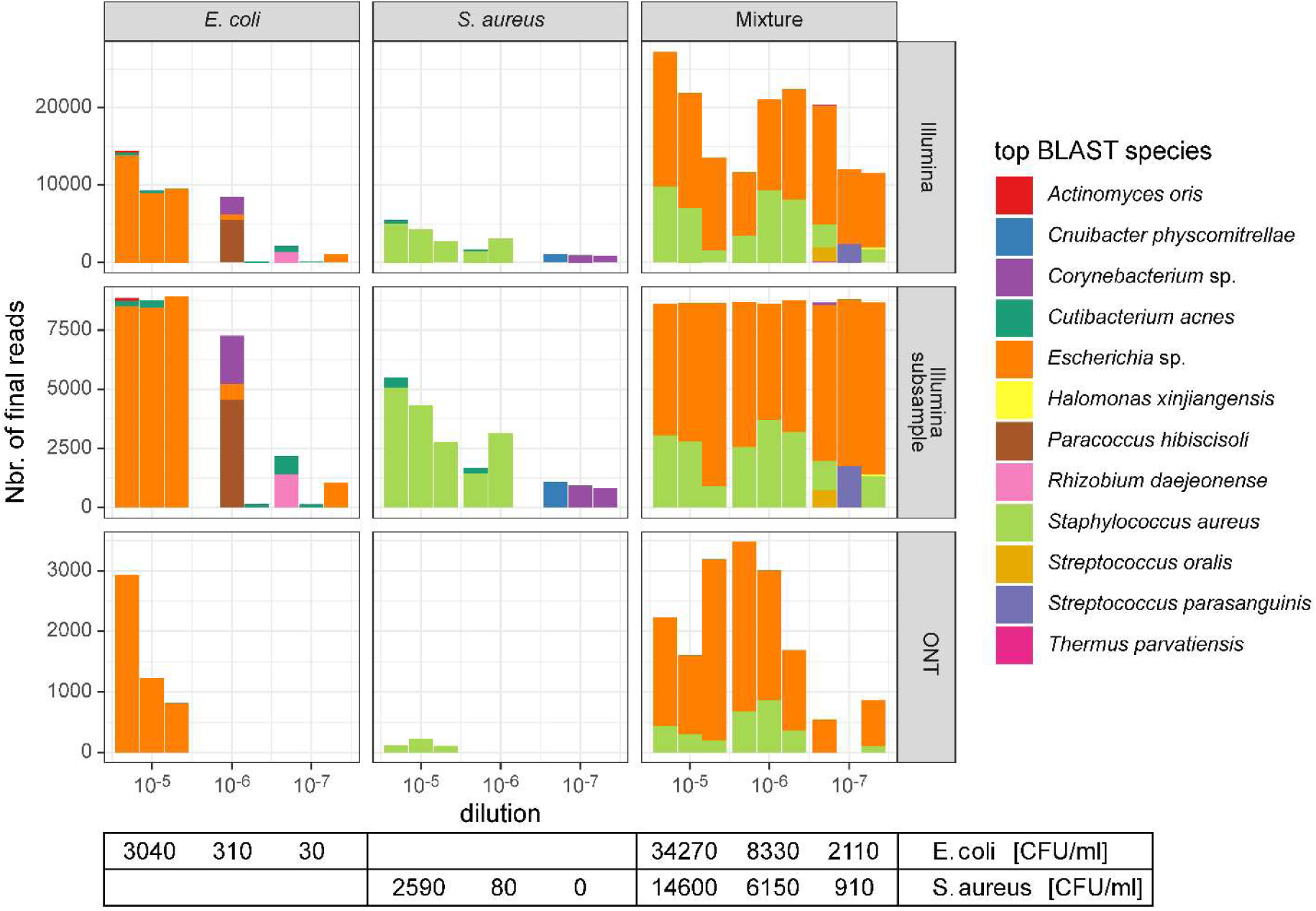
Sensitivity and specificity based on dilutions series of pure bacterial liquid cultures. Serial dilutions for *E. coli*, *S. aureus* and a mixture of both were prepared and subjected to our laboratory and bioinformatic workflows (**Figure 1**). Reads assigned to *Escherichia* sp. and *Shigella* sp. were grouped as *Escherichia* sp. due to their high similarity of the 16S rRNA gene across those species. Similarly, *Corynebacterium* species were grouped together. Species associated with less than 100 reads in one sample are not displayed. The observed number of CFUs (colony forming units) per ml of the diluted liquid cultures are indicated at the bottom of the figure.

### Clinical samples

We analyzed 101 clinical samples that were routinely collected over a 1.5-year period, from patients who gave their general consent, and for which the 16S PCR amplification was successful (> 1 ng/µl DNA concentration). These samples represented a diverse range of clinical materials, encompassing 43 distinct sample types, and were predominantly derived from joint and spine biopsies (**Supplementary Table 3**). All samples underwent sequencing using both ONT and Illumina on the same DNA extracts. Across both sequencing methods, a total of 167 species were detected with at least 100 assigned reads, and 115 (68.9%) of these species were shared between the two methods (**Figure 3A, Supplementary Table 4**), while 23 (13.8%) species were uniquely detected by ONT and 29 (17.4%) species were exclusively found by Illumina. Subsampling the Illumina reads reduced the number of detected species, resulting in only 109 (65.3%) shared species and 5 (3.0%) species unique to Illumina. The distribution of species across samples was comparable between the two methods. ONT detected a species in an average of 2.23 ± 2.97 samples, while Illumina detected a species in an average of 2.08 ± 3.11 samples (2.18 ± 3.10 for subsampled Illumina data; **Figure 3B**). *Cutibacterium acnes* was the most common identified species (in 20% and 30% of the samples, based on ONT and Illumina sequencing, respectively), with average relative abundances of 51 ± 40% with ONT, and 39 ± 41% with Illumina sequencing, across samples it was present in. Other common species belonged to *Staphylococcus* and *Streptococcus* genera. We found that the overall species detection pattern was consistent between the two methods when considering the relative abundances of species within each sample (**Figure 3B**). Species uniquely detected by either Illumina or ONT generally had lower average relative abundances (3% on average). Aggregating the abundances of all species across all samples further showed that the detection patterns for Illumina and ONT were highly similar (**Figure 3C**), and subsampling the Illumina reads did not significantly alter these trends. Consistent with the prevalence data, *C. acnes* and several *Staphylococcus* and *Streptococcus* species were highly represented in the total dataset (**Figure 3C**). As an estimate of bacterial diversity per sample, the average number of species per sample was 3.0 ± 2.7 for ONT, and 3.0 ± 3.4 for Illumina (2.5 ± 2.3 for the subsampled Illumina data). Subsampling the Illumina reads significantly reduced the number of species per sample (paired *t* test, p-value < 0.001), possibly due to a reduction of the number of detected contaminants. To quantitatively evaluate the differences between the two methods, we calculated the overlap of species for each sample: On average, 97.1 ± 5.9%, of species detected by ONT (weighted by their relative abundance) were also found by Illumina, while 98.7 ± 2.8% of Illumina species (weighted by their relative abundance) were also detected by ONT. The overall congruence of relative abundance of shared species between methods (calculated by averaging the sum of the minimum relative abundances of the co-detected species across all samples) was 93.5 ± 7.6%. To assess whether the sequencing platform significantly influenced the species composition, we performed a Permutational Multivariate Analysis of Variance (PERMANOVA). This multivariate approach accounts for the collective variation across all taxa by evaluating the total dissimilarity between samples (18). Using either Bray-Curtis distances for relative abundance and Jaccard distances for presence-absence data, we found no significant differences in species composition between the two platforms. Lastly, a comparison of consensus sequence accuracy based on BLAST identity showed that sequences had an average sequence identity to the reference of 99.88 ± 0.39 % (weighted by reads) when generated by ONT, and of 99.83 ± 0.42 % with Illumina. Despite achieving statistical significance (p<0.001; paired *t* test weighted by read number), this marginal difference indicates that both platforms provided near-identical consensus accuracy.

**Figure 3.**
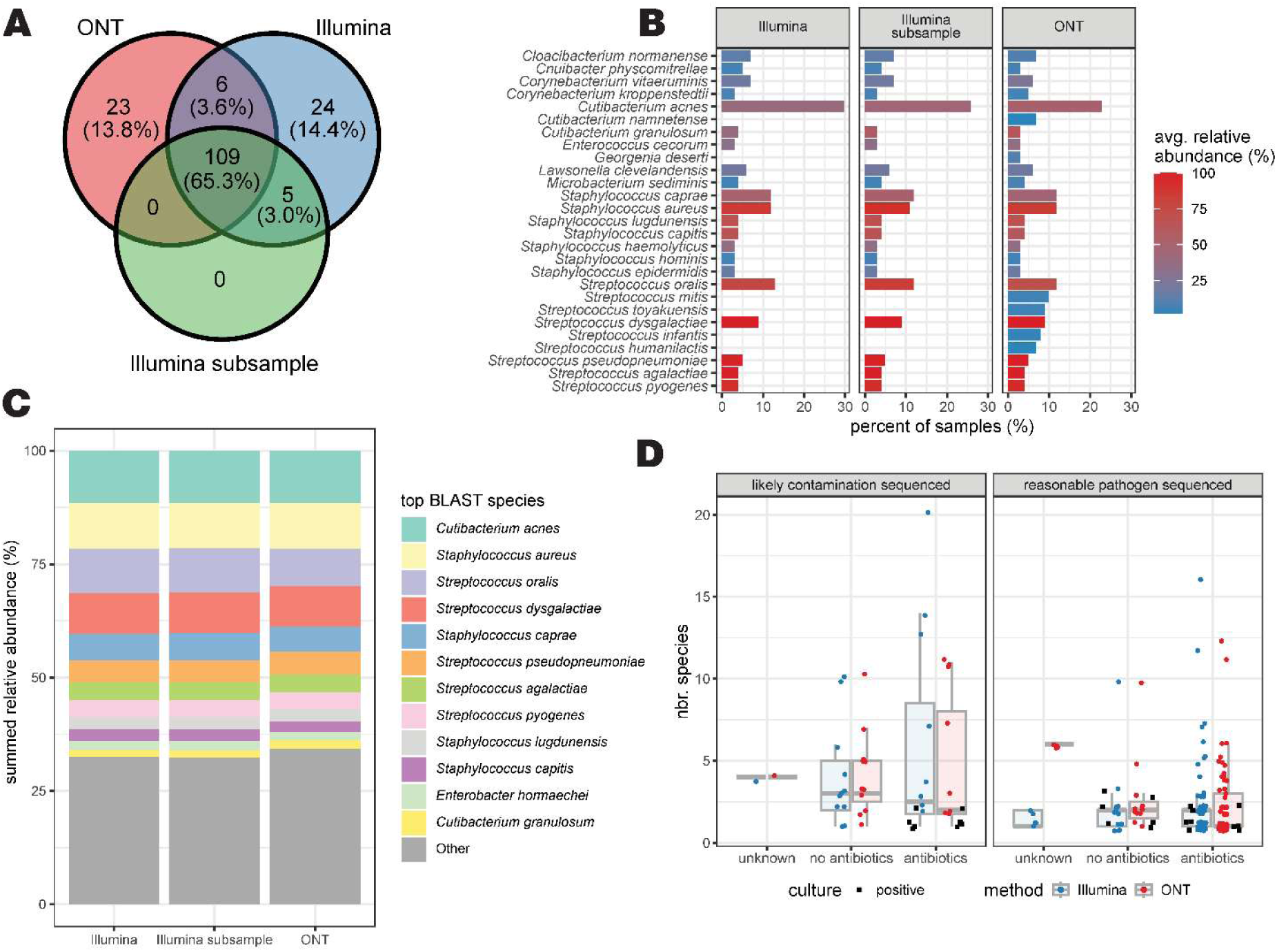
Detected bacterial species in routine clinical samples. **A)** Venn-diagram of the number of species shared between sequencing methods in clinical samples (n=101). In parentheses the percentages of all the species across samples are given for each case (167 detected species in total). **B)** Species prevalence (i.e. percent of samples in which a species is present) across sequencing technologies. The average relative abundance across the samples in which a species was found present is indicated by a color range (red = 100%, blue = 0%). Only species that are present in at least 3 samples in at least one method are depicted. **C)** Barplots of the summed relative abundances of detected species across all samples for each of the sequencing methods. Shown are the 12 most abundant species, the remaining are grouped into the category “Other”. **D)** Boxplot of the number of species identified by sequencing method, grouped by antibiotic pre-treatment of the samples and by the results of the clinical assessment. Each dot represents an individual sample. The shape of the dot indicates the culture result for that specimen, regardless of concordance with sequencing data. No significant difference was observed between antibiotic treatments for either method. The number of species was significantly higher in samples classified as likely contamination compared to those containing a reasonable pathogen for both lllumina (p=0.02) and ONT (p=0.03) workflows.

### Clinical plausibility of the results

Sequencing results were first compared to the routine culturing results. For 12 samples a positive culture from the same specimen was obtained, while for 86 samples no growth was detected **(Figure 3D; Supplementary Table 3)**. For 8 of the culture-positive samples, species identification agreed with both sequencing workflows. In three of the discordant cases, in which culture and sequencing results did not match, *C. albicans* was detected by culture while sequencing only detected *S. caprae* and *S. haemolyticus* (all three samples were from the same patient). It is not surprising that the sequencing did not detect *C. ablicans* since the 16S primers are prokaryote specific (7). For the other discordant case, culturing only detected a likely contamination by *Micrococcus* sp., while sequencing only detected contamination by *Nesterenkonia* sp. Overall, using the culture results as reference for valid species detection, we can calculate a positive predictive value (PPV; TP/(TP+FP)) of 0.08% for the NGS sequencing in general. The low PPV stems mainly from the high number of culture negative samples.

To further evaluate the clinical plausibility of the NGS findings, an experienced infectiologist retrospectively reviewed all sequencing results obtained for the clinical samples with respect to patient medical history and complementary microbiological results. The clinical review found a high level of agreement between the two sequencing methods. Overall, in 77 (76.2%) samples a likely pathogen was observed, while the data for the remaining 24 (23.8%) samples suggested the detection of likely contaminations. Importantly, there was no instance of discordant classification (pathogen vs. contamination) between the two sequencing methods for any sample. This means that, if we combine the clinical review and the presence of a positive culture as a composite reference for a true detection, a PPV of 76.2% was achieved by both ONT and Illumina sequencing workflows. We further assessed diagnostic agreement using weighted Cohen’s *kappa* based on three categories: Identification of pathogen only, pathogen plus contamination, and contamination only (**Supplementary Table 5**). Using the respective raters “ONT plus clinical review” and “Illumina plus clinical review”, we obtained a *kappa* of 0.81 ± 0.04 with the full Illumina dataset, which increased to 0.89 ± 0.03 when using the subsampled Illumina dataset.

We also observed that, for both sequencing platforms, samples considered contaminated contained significantly more detected species per sample than those with a likely pathogen (Illumina: 4.96 ± 4.97 vs 2.36 ± 2.54, p = 0.02; ONT: 4.33 ± 3.40 vs 2.66 ± 2.38, p = 0.03; **Figure 3D)**. Furthermore, we assessed the potential impact of patient antibiotic treatment prior to sampling and sequencing, which was documented in 94% of the samples (69 samples with treatment, 26 samples without). There was no significant correlation between antibiotic pretreatment and the number of species detected for either sequencing workflow. Interestingly, in samples with antibiotic pretreatment significantly more pathogens than contaminations were detected (Fisher exact test, p-value = 0.02; **Supplementary Tables 3 and 4**).

### Cost and turnaround time estimation

We estimated that consumables for a batch size of 24 samples processed on a single flow cell resulted in approximate per-sample costs of 35 CHF (US$44) for ONT and 155 CHF (US$200) for Illumina. The effective price difference is likely to be even greater in routine diagnostic settings, as batch sizes are often smaller than 24 samples. This is primarily because ONT flow cells can be reused if only a small number of samples is sequenced, while Illumina sequencing cartridges are single-use, regardless of the number of sequenced samples. To estimate the turnaround time of the two sequencing approaches, we considered the time required for library preparation, sequencing, and bioinformatic analysis (including data transfer, quality control, and report generation) for a 24-sample batch. The ONT workflow required approximately 5, 2, and 0.5 h for each of the steps, respectively. In contrast, the Illumina workflow required 7, 43, and 0.5 h, respectively. These estimates for the bioinformatic component rely on the implementation of established and automated analysis pipelines. These findings underscore the ONT-based workflow’s advantage in speed, demonstrating a much faster turnaround than the Illumina-based workflow.

## Discussion

This study demonstrates that ONT sequencing provides a time- and cost-efficient approach for 16S rRNA sequencing of low-diversity samples under routine clinical diagnostic conditions. Using a controlled dilution series and a large panel of 101 routine clinical samples, we showed that ONT 16S rRNA gene sequencing achieves a level of sequencing accuracy that is comparable to that of the more established Illumina sequencing method. In 76% of clinical samples, the combination of an IVD certified 16S amplification protocol together with ONT sequencing successfully identified a likely human pathogen, as confirmed by clinical review of the cases. Furthermore, for batch sizes of around 24 samples, ONT was much more time and cost efficient than the Illumina-based workflow.

In our verification experiments, the limit of detection achieved using the ONT platform was in the range of 100 to 1,000 CFUs/ml (**Figure 2**). In a recent study standardizing 16S rRNA gene sequencing for culture-negative infections (19), the authors evaluated ONT analytical sensitivity using DNA genome copies per µl, noting that species detection varied, and observed better sensitivity with the MinION flow cell than with Flongle, potentially due to differences in sequencing depth. Another investigation that focused on direct bacterial identification in sterile body fluids estimated a slightly lower detection limit of 90 CFUs/ml (20). This difference is likely attributable to their substantially higher sequencing depth, reporting approximately ten times more reads per sample than generated in our study. In comparison, Illumina sequencing demonstrated a slightly lower detection limit in our controlled dilution series (**Figure 2**), irrespective of whether the full or a subsampled dataset was analyzed. This aligns with findings from other 16S studies, where Illumina sequencing was observed to capture more low abundant species, and thus greater species richness in respiratory microbial communities (21). The slightly higher detection limit of ONT could potentially lead to higher numbers of false negatives and in the presence of contaminants might lead to misdetection, which would increase the number of false positives. For our clinical samples the rate of false negatives is controlled by the 16S PCR amplification protocol, because samples with a negative RT-PCR result are not further subjected to sequencing. As the sensitivity of the 16S amplification protocol has been previously established (7), we focused primarily on comparing the sequencing methodologies rather than re-evaluating false negative rates. Notably, regarding false positives in clinical samples, we observed no difference in incidence between Illumina and ONT sequencing.

An interesting finding from the controlled dilution experiments was the detection of more contamination-associated reads with Illumina than with ONT sequencing, when a minimum threshold of 100 reads per species for identification was set **(Figure 2)**. However, when examining routine clinical samples, both sequencing methods appeared equally likely to detect non-pathogenic contaminants **(Figure 3D)**, which is reflected in the Cohen’s *kappa* of 0.81 ± 0.04. The correct interpretation and reporting of such low-abundance reads is critical for clinical decision making. One study on mock communities found that ONT displayed remarkably high accuracy compared to Illumina sequencing (22), though the use of full-length 16S for ONT and shorter amplicons for Illumina in that study represents a confounding factor. To address the “tail” of low-abundant species (often contaminants) observed in sequencing data, prior studies have implemented relative abundance filters, such as filtering out species below 0.1% (23) or 0.06% (20). While filtering by relative abundance may work well for monomicrobial samples, it was shown to fail for polymicrobial samples (20), underscoring the complexity of robustly differentiating true pathogens from contaminants in complex clinical scenarios. We found *Cutibacterium acnes* to be one of the most common species in the investigated clinical samples **(Figure 3B)**. Because *C. acnes* is a skin commensal and can also be an opportunistic pathogen, differentiating a true infection from a contamination can be challenging (24). In eleven (11%) of our clinical samples *C. acnes* was the likely pathogen as evaluated by the clinical review, while in the remaining 10-20 samples, a potential implication of *C. acnes* as a pathogen could not be unambiguously determined (**Supplementary Tables 3-4**). In those latter cases, *C. acnes* presence might thus represent a contamination occurring during the sampling or laboratory processes. The high occurrence of *C. acnes* may also be attributable to the significant proportion of joint biopsies among our clinical samples. *C. acnes* has gained notoriety for its role in implant-associated infections and has also been recognized as a significant pathogen in cerebrovascular infections, breast implant fibrosis, and cardiovascular device infections (24).

Overall, we observed an average congruence in species identification of 93.5% (weighted by abundance) between the two sequencing workflows. This direct comparison, conducted on a large set of routine clinical specimens prospectively collected over 18 months, confirms that ONT sequencing is equally effective and accurate as Illumina sequencing, for clinical pathogen diagnostics. Furthermore, the ONT platform in combination with the LORCAN bioinformatic pipeline demonstrated a slightly higher consensus sequence accuracy (99.88 ± 0.39% sequence identity to reference) compared to sequences generated by Illumina and DADA2 (15) (99.83 ± 0.42%). While several studies have explored the feasibility of ONT 16S rRNA gene sequencing in diagnostics, primarily comparing results to culture (20, 25–28), MALDI-TOF (29, 30), or Sanger sequencing (19, 30–32), there is a notable absence of studies directly comparing ONT versus Illumina sequencing using a large cohort of clinical samples. A small scale study found that the microbial composition determined with Illumina V3-V4 sequencing closely matched to the one determined with ONT full-length sequencing in tracheal aspirates samples (23), as well as in a mock community. Similarly, an older study in 2019 showed both technologies could accurately identify bacterial genera in a well-characterized reference sample (22). The observed low PPV of both sequencing workflows relative to culture is primarily driven by the high prevalence of culture-negative results among the specimens. This discrepancy is anticipated, as conventional culturing frequently underperforms when identifying low-abundance or fastidious organisms (2). Notably, previous literature demonstrated that 16S sequencing offers superior sensitivity for pathogen detection; for instance, Egli et al. (7) reported that 36.5% of PCR-positive samples remained culture-negative, a finding supported by clinical correlation from independent patient specimens. This diagnostic gap represents a clear clinical benefit, as corroborated by several studies highlighting the increased yield of 16S sequencing over traditional methods (20, 25–28). Furthermore, it is important to note that both ONT and Illumina workflows exhibited equivalent PPV, regardless of whether culture alone or a composite reference (incorporating clinical review) was utilized as the benchmark.

The decision to use the V3-V4 targeting IVD-R kit in our study was driven by its analytical robustness and established clinical validation: The kit provides broad-range detection of >345 bacterial species of clinical relevance (33) and demonstrates high concordance with traditional culture results (7). Furthermore, relying on a standardized IVD-R certified kit guarantees reproducibility and rigorous quality control. Transitioning to full-length 16S diagnostics in the future will depend on the commercial availability of IVD-R certified, DNA-free reagent kits capable of full-length amplification. We acknowledge that targeting only the V3-V4 region restricts taxonomic resolution; full-length 16S sequencing generally yields superior species-level discrimination, such as distinguishing *E. coli* from *Shigella* (20) or differentiating among *Staphylococcus* species (23). From a technical standpoint, shorter amplicons typically offer superior PCR amplification efficiency, especially in clinical samples containing fragmented DNA (32). From a clinical decision standpoint, the advantages of species-level versus genus-level identification are not always clear-cut, and are highly dependent on the specific genus. For instance, some evidence suggests that species-level identification of coagulase-negative staphylococci in blood cultures may not significantly impact clinical management with regard to antibiotic utilization, length of hospital stay or mortality (34). Other studies underscore the clinical importance of species-level identification, demonstrating that flucloxacillin resistance and associated mortality rates vary significantly across different species (35).

In conclusion, untargeted 16S ONT sequencing coupled with IVD-R PCR kit holds a very positive outlook in clinical diagnostics of low-diversity samples, primarily due to its proven advantages in shorter turnaround time and cost efficiency, especially with small batch sizes. The added benefit of 16S sequencing to culture has been well-documented, and the successful demonstration that ONT sequencing achieves an analytical accuracy comparable to that of conventional Illumina sequencing, confirms its viability as a mainstream clinical tool. Future efforts should prioritize maximizing the platform’s utility by addressing current limitations. Specifically, using longer sequences or complementary genetic targets could be crucial to enhance taxonomic resolution (36), especially in combination with full-length curated databases of 16S rRNA genes (37–39). Finally, continued development is necessary for robust bioinformatic strategies to accurately interpret low-abundance reads, which represent contaminants, especially in complex clinical scenarios where filtering methods successful for monomicrobial samples may fail for polymicrobial samples.

## Data Availability

Raw sequencing data is available under the BioProject accession number: PRJEB102560. Supplementary tables can be accessed under: https://zenodo.org/records/17811713.

