## Supplementary Figure 1; Supplementary Tables 1-5 for "Performance and Practicality of 16S Nanopore Sequencing for Routine Bacterial Identification in Clinical Samples"

### 1 Supplementary Figure 1

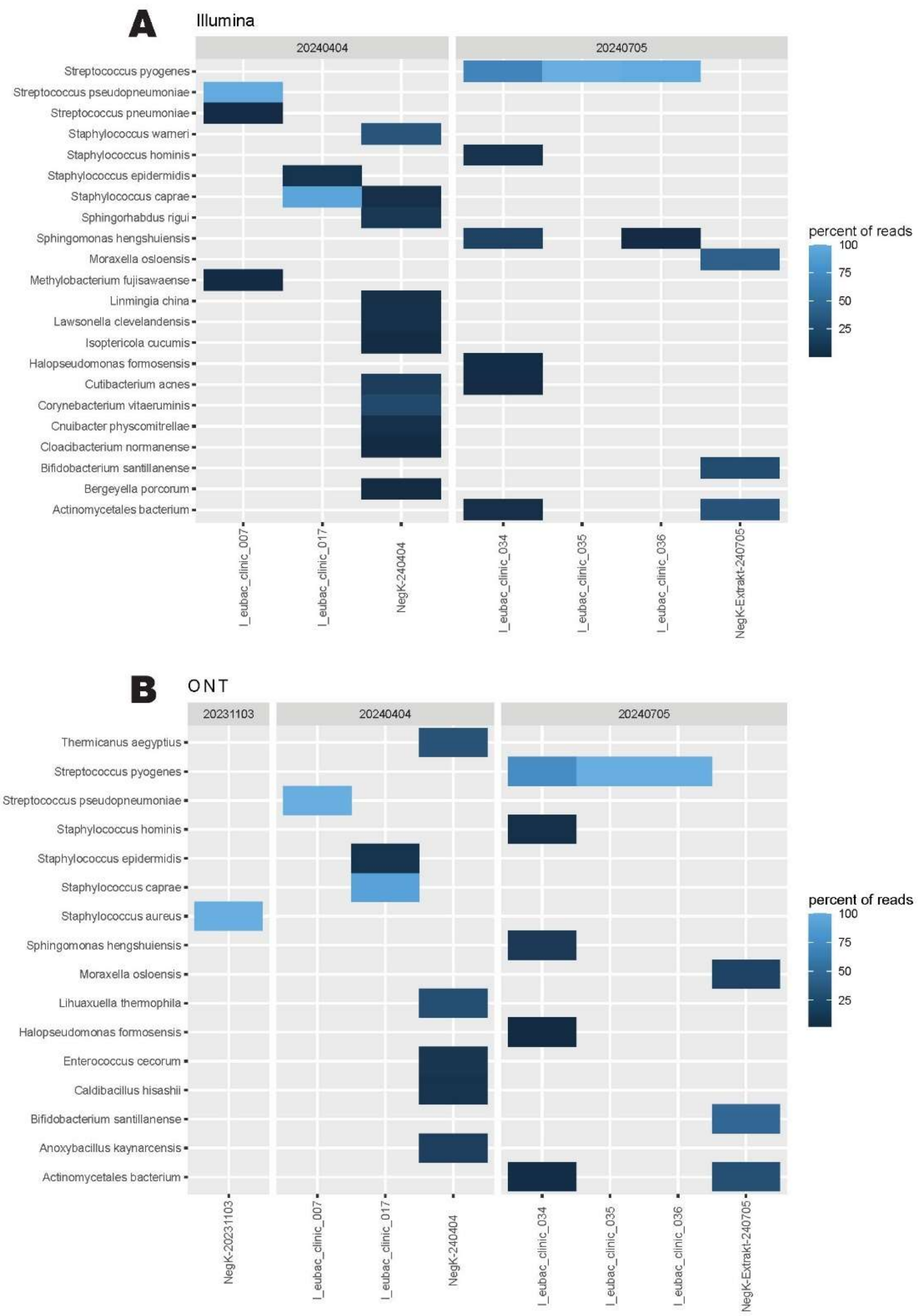

**Supplementary Figure 1. Heatmap of species relative abundance in the negative controls and their corresponding samples**. The relative abundances (color ranging from 0% = black to 100%= bright blue) in each of the negative controls and their corresponding samples grouped by processing batch, are displayed for both of the sequencing methods Illumina **(A)** and ONT **(B)**.

### 2 Supplementary Tables

Supplementary tables can be accessed under: https://zenodo.org/records/17811713.

**Supplementary Table 1.** Read numbers for both ONT and MiSeq sequencing, before and after filtering.

| Sample_ID | Nbr. Species assinged reads Illumina | Nbr. Species assigned reads Illumina subsample | Nbr. species assigned reads ONT | Nbr. Illumina filtered reads | Nbr. Illumina raw reads | Nbr. ONT filtered reads | Nbr. ONT raw reads |
|---|---|---|---|---|---|---|---|
| I_eubac_clinic_001 | 40375 | 8229 | 5226 | 44075 | 363,153 | 5,444 | 5,758 |
| I_eubac_clinic_002 | 42972 | 8249 | 6949 | 47089 | 265,570 | 7,348 | 7,937 |
| I_eubac_clinic_003 | 20079 | 8289 | 3941 | 21900 | 203,717 | 4,055 | 4,350 |
| I_eubac_clinic_004 | 21530 | 3772 | 8168 | 53311 | 221,645 | 9,000 | 23,836 |
| I_eubac_clinic_005 | 23832 | 6024 | 8723 | 50188 | 180,989 | 9,000 | 43,301 |
| I_eubac_clinic_006 | 22984 | 4410 | 8822 | 46562 | 205,001 | 9,000 | 69,076 |
| I_eubac_clinic_007 | 58757 | 8512 | 8633 | 60106 | 168,376 | 9,000 | 18,551 |
| I_eubac_clinic_008 | 45174 | 8738 | 8908 | 46173 | 172,809 | 9,000 | 16,338 |
| I_eubac_clinic_009 | 37745 | 8752 | 7854 | 39542 | 178,118 | 7,920 | 8,255 |
| I_eubac_clinic_010 | 26070 | 8809 | 8913 | 26430 | 167,675 | 9,000 | 12,620 |
| I_eubac_clinic_011 | 46282 | 8722 | 8997 | 48083 | 187,677 | 9,000 | 9,631 |
| I_eubac_clinic_012 | 40639 | 8775 | 8996 | 41678 | 165,139 | 9,000 | 13,648 |
| I_eubac_clinic_013 | 47300 | 8857 | 6902 | 47910 | 361,187 | 7,061 | 7,245 |
| I_eubac_clinic_014 | 47532 | 8799 | 8887 | 48168 | 243,235 | 9,000 | 11,558 |
| I_eubac_clinic_015 | 28972 | 8756 | 2090 | 29932 | 292,635 | 2,177 | 2,268 |
| I_eubac_clinic_016 | 44726 | 8833 | 8903 | 45526 | 133,200 | 9,000 | 12,424 |
| I_eubac_clinic_017 | 58663 | 8786 | 6240 | 62897 | 273,828 | 6,482 | 6,670 |
| I_eubac_clinic_018 | 32024 | 8648 | 4292 | 33505 | 132,918 | 4,444 | 4,864 |
| I_eubac_clinic_019 | 24551 | 8368 | 1127 | 26282 | 354,219 | 1,444 | 1,526 |
| I_eubac_clinic_020 | 39782 | 6763 | 8799 | 53142 | 189,850 | 9,000 | 19,437 |
| I_eubac_clinic_021 | 32141 | 8165 | 8884 | 35781 | 169,118 | 9,000 | 14,614 |
| I_eubac_clinic_022 | 32223 | 7853 | 8973 | 36674 | 188,658 | 9,000 | 18,888 |
| I_eubac_clinic_023 | 52894 | 8568 | 8738 | 56607 | 152,477 | 9,000 | 23,151 |
| I_eubac_clinic_024 | 50663 | 8863 | 8749 | 52732 | 154,997 | 9,000 | 27,477 |
| I_eubac_clinic_025 | 2363 | 2364 | 6589 | 2713 | 46,081 | 6,833 | 7,030 |
| I_eubac_clinic_026 | 9621 | 8762 | 6832 | 9888 | 65,866 | 6,983 | 7,200 |
| I_eubac_clinic_027 | 4404 | 4406 | 2271 | 4572 | 24,147 | 2,503 | 2,673 |
| I_eubac_clinic_028 | 30172 | 8335 | 4605 | 32697 | 222,394 | 4,910 | 5,097 |
| I_eubac_clinic_029 | 49629 | 8833 | 8715 | 50405 | 128,331 | 9,000 | 39,311 |
| I_eubac_clinic_030 | 44348 | 8795 | 8685 | 45064 | 126,872 | 9,000 | 50,970 |
| I_eubac_clinic_031 | 41495 | 8891 | 7159 | 42010 | 134,581 | 7,380 | 7,594 |
| I_eubac_clinic_032 | 38912 | 8896 | 5663 | 39327 | 159,161 | 5,896 | 6,031 |
| I_eubac_clinic_033 | 48976 | 8675 | 8554 | 49505 | 158,343 | 9,000 | 38,340 |
| I_eubac_clinic_034 | 42735 | 8738 | 6460 | 43991 | 202,324 | 6,740 | 6,921 |
| I_eubac_clinic_035 | 32086 | 8865 | 5012 | 32604 | 164,782 | 5,014 | 5,127 |

| | | | | | | | |
|---|---|---|---|---|---|---|---|
| l_eubac_clinic_036 | 45224 | 8718 | 8951 | 46470 | 138,441 | 9,000 | 23,923 |
| l_eubac_clinic_037 | 31767 | 8912 | 8845 | 32867 | 145,057 | 9,000 | 17,545 |
| l_eubac_clinic_038 | 8249 | 8251 | 1056 | 8944 | 90,829 | 1,356 | 1,428 |
| l_eubac_clinic_039 | 27409 | 8295 | 4386 | 28393 | 130,519 | 4,970 | 5,159 |
| l_eubac_clinic_040 | 24587 | 8828 | 1257 | 25155 | 157,200 | 1,353 | 1,405 |
| l_eubac_clinic_041 | 43590 | 8766 | 8677 | 44654 | 127,708 | 9,000 | 15,165 |
| l_eubac_clinic_042 | 22686 | 8875 | 4308 | 23019 | 84,387 | 4,309 | 4,505 |
| l_eubac_clinic_043 | 27716 | 8896 | 8020 | 28022 | 114,832 | 8,026 | 8,270 |
| l_eubac_clinic_044 | 24622 | 8885 | 6819 | 24971 | 80,921 | 6,821 | 7,082 |
| l_eubac_clinic_045 | 47698 | 7437 | 8701 | 66879 | 135,597 | 9,000 | 51,226 |
| l_eubac_clinic_046 | 21955 | 4755 | 736 | 35314 | 202,341 | 807 | 920 |
| l_eubac_clinic_047 | 46121 | 8209 | 8400 | 51480 | 135,126 | 9,000 | 10,919 |
| l_eubac_clinic_048 | 42846 | 8636 | 6887 | 45341 | 152,995 | 7,146 | 7,392 |
| l_eubac_clinic_049 | 42924 | 8822 | 1844 | 44344 | 161,576 | 1,984 | 2,079 |
| l_eubac_clinic_050 | 21153 | 8709 | 8399 | 25024 | 184,302 | 9,000 | 17,229 |
| l_eubac_clinic_051 | 6987 | 6987 | 208 | 7698 | 270,405 | 227 | 276 |
| l_eubac_clinic_052 | 13579 | 8549 | 207 | 14412 | 130,958 | 251 | 305 |
| l_eubac_clinic_053 | 24383 | 7729 | 4863 | 33363 | 204,930 | 5,380 | 5,700 |
| l_eubac_clinic_054 | 28530 | 7300 | 8989 | 37131 | 193,723 | 9,000 | 11,485 |
| l_eubac_clinic_055 | 16062 | 8498 | 2120 | 18533 | 293,802 | 2,233 | 2,299 |
| l_eubac_clinic_056 | 19718 | 8489 | 1198 | 23335 | 179,763 | 1,256 | 1,299 |
| l_eubac_clinic_057 | 13756 | 5522 | 8946 | 25075 | 142,837 | 9,000 | 13,864 |
| l_eubac_clinic_058 | 12952 | 8770 | 1597 | 13438 | 261,129 | 1,851 | 1,892 |
| l_eubac_clinic_059 | 21885 | 7016 | 8923 | 29223 | 117,618 | 9,000 | 14,791 |
| l_eubac_clinic_060 | 2652 | 2652 | 313 | 3220 | 98,535 | 339 | 411 |
| l_eubac_clinic_061 | 24416 | 8342 | 1277 | 28174 | 221,245 | 1,371 | 1,499 |
| l_eubac_clinic_062 | 18526 | 8712 | 1731 | 20444 | 223,596 | 1,808 | 1,879 |
| l_eubac_clinic_063 | 3565 | 3565 | 1888 | 4248 | 119,615 | 2,212 | 2,297 |
| l_eubac_clinic_064 | 2518 | 2517 | 5695 | 2853 | 118,434 | 5,864 | 6,346 |
| l_eubac_clinic_065 | 20372 | 8820 | 8857 | 24757 | 129,712 | 9,000 | 23,633 |
| l_eubac_clinic_066 | 9100 | 8794 | 8799 | 9318 | 93,853 | 9,000 | 16,930 |
| l_eubac_clinic_067 | 10613 | 7854 | 1241 | 12213 | 168,067 | 1,450 | 1,574 |
| l_eubac_clinic_068 | 21622 | 8014 | 6595 | 24381 | 165,554 | 7,439 | 7,771 |
| l_eubac_clinic_069 | 25031 | 7192 | 6914 | 28542 | 120,917 | 8,753 | 9,295 |
| l_eubac_clinic_070 | 8521 | 7616 | 898 | 10109 | 126,452 | 929 | 978 |
| l_eubac_clinic_071 | 2362 | 2364 | 395 | 2506 | 67,994 | 463 | 497 |
| l_eubac_clinic_072 | 14387 | 6474 | 6595 | 18916 | 73,201 | 9,000 | 12,993 |
| l_eubac_clinic_073 | 11811 | 7942 | 7092 | 13277 | 106,667 | 7,309 | 7,852 |
| l_eubac_clinic_074 | 15303 | 8695 | 2201 | 16010 | 94,254 | 2,585 | 2,650 |
| l_eubac_clinic_075 | 14181 | 8049 | 7510 | 18011 | 57,746 | 7,841 | 7,986 |
| l_eubac_clinic_076 | 12531 | 7497 | 2180 | 14468 | 88,812 | 2,514 | 2,928 |
| l_eubac_clinic_077 | 20718 | 6944 | 3847 | 26780 | 97,735 | 4,060 | 4,213 |
| l_eubac_clinic_078 | 13491 | 7176 | 3550 | 16903 | 61,541 | 3,585 | 3,801 |
| l_eubac_clinic_079 | 7871 | 7871 | 244 | 8118 | 131,756 | 318 | 375 |
| l_eubac_clinic_080 | 14826 | 6093 | 8332 | 31013 | 104,178 | 9,000 | 11,472 |
| l_eubac_clinic_081 | 9006 | 7693 | 8995 | 10553 | 55,319 | 9,000 | 15,506 |

| | | | | | | | |
|---|---|---|---|---|---|---|---|
| l_eubac_clinic_082 | 12826 | 7143 | 4792 | 15973 | 79,169 | 4,863 | 5,196 |
| l_eubac_clinic_083 | 2382 | 2383 | 169 | 2534 | 65,862 | 314 | 342 |
| l_eubac_clinic_084 | 7819 | 7700 | 3633 | 8448 | 74,166 | 4,376 | 4,636 |
| l_eubac_clinic_085 | 2728 | 2728 | 654 | 3049 | 70,122 | 788 | 881 |
| l_eubac_clinic_086 | 700 | 700 | 232 | 739 | 22,935 | 239 | 261 |
| l_eubac_clinic_087 | 1604 | 955 | 1384 | 14892 | 225,313 | 9,000 | 87,505 |
| l_eubac_clinic_088 | 8039 | 4777 | 4369 | 15126 | 980,641 | 4,577 | 8,500 |
| l_eubac_clinic_089 | 3628 | 2028 | 2381 | 16258 | 222,600 | 7,183 | 36,543 |
| l_eubac_clinic_090 | 25037 | 7315 | 8135 | 30259 | 401,163 | 9,000 | 81,479 |
| l_eubac_clinic_091 | 18775 | 7978 | 8546 | 21029 | 644,886 | 9,000 | 27,909 |
| l_eubac_clinic_092 | 4424 | 3313 | 3560 | 11939 | 588,224 | 7,942 | 29,589 |
| l_eubac_clinic_093 | 19305 | 6483 | 7987 | 25989 | 541,134 | 9,000 | 51,028 |
| l_eubac_clinic_094 | 6465 | 6467 | 8582 | 7069 | 816,217 | 9,000 | 19,475 |
| l_eubac_clinic_095 | 29252 | 8109 | 8745 | 36970 | 551,601 | 9,000 | 39,855 |
| l_eubac_clinic_096 | 6263 | 3577 | 2583 | 15720 | 838,463 | 4,017 | 13,176 |
| l_eubac_clinic_097 | 27206 | 7143 | 7517 | 30383 | 599,745 | 9,000 | 16,330 |
| l_eubac_clinic_098 | 16433 | 2813 | 8711 | 55850 | 357,065 | 9,000 | 59,206 |
| l_eubac_clinic_099 | 37683 | 7702 | 8682 | 44809 | 152,098 | 9,000 | 112,070 |
| l_eubac_clinic_100 | 36400 | 8710 | 8703 | 37373 | 201,021 | 9,000 | 24,530 |
| l_eubac_clinic_101 | 41395 | 8618 | 8768 | 43999 | 376,900 | 9,000 | 21,704 |
| 1-eco5-1 | 14418 | 8857 | 2928 | 15192 | 133,716 | 3,310 | 3,434 |
| 10-sau5-1 | 5500 | 5501 | 123 | 5718 | 227,667 | 151 | 164 |
| 11-sau5-2 | 4317 | 4317 | 223 | 4403 | 169,920 | 236 | 259 |
| 12-sau5-3 | 2776 | 2774 | 112 | 3172 | 205,286 | 121 | 134 |
| 13-sau6-1 | 1688 | 1688 | 0 | 1913 | 308,844 | 116 | 134 |
| 14-sau6-2 | 3142 | 3141 | 0 | 3184 | 359,979 | 68 | 100 |
| 16-sau7-1 | 1090 | 1090 | 0 | 1155 | 289,781 | 72 | 92 |
| 17-sau7-2 | 935 | 935 | 0 | 1324 | 111,245 | 209 | 246 |
| 18-sau7-3 | 830 | 830 | 0 | 874 | 104,737 | 111 | 131 |
| 19-mix5-1 | 27246 | 8605 | 2231 | 29597 | 151,310 | 2,407 | 2,545 |
| 2-eco5-2 | 9261 | 8771 | 1234 | 9373 | 108,352 | 1,579 | 1,644 |
| 20-mix5-2 | 21912 | 8649 | 1608 | 23929 | 132,640 | 1,836 | 1,967 |
| 21-mix5-3 | 13539 | 8634 | 3195 | 15254 | 91,218 | 3,503 | 3,656 |
| 22-mix6-1 | 11726 | 8680 | 3482 | 12156 | 112,799 | 3,740 | 3,905 |
| 23-mix6-2 | 21048 | 8591 | 3016 | 22615 | 119,396 | 3,261 | 3,497 |
| 24-mix6-3 | 22402 | 8766 | 1693 | 23174 | 116,360 | 1,917 | 2,028 |
| 25-mix7-1 | 20404 | 8665 | 550 | 21988 | 213,937 | 816 | 874 |
| 26-mix7-2 | 12043 | 8821 | 0 | 12281 | 220,425 | 323 | 353 |
| 27-mix7-3 | 11539 | 8670 | 864 | 12017 | 252,523 | 1,104 | 1,191 |
| 3-eco5-3 | 9527 | 8912 | 822 | 9618 | 101,227 | 1,012 | 1,065 |
| 5-eco6-2 | 8512 | 7259 | 0 | 9959 | 258,910 | 162 | 184 |
| 6-eco6-3 | 165 | 165 | 0 | 181 | 154,962 | 47 | 65 |
| 7-eco7-1 | 2192 | 2192 | 0 | 2362 | 101,123 | 60 | 79 |
| 8-eco7-2 | 142 | 142 | 0 | 288 | 315,184 | 41 | 57 |
| 9-eco7-3 | 1050 | 1050 | 0 | 1356 | 185,249 | 85 | 109 |
| NegK-20231103 | 0 | 0 | 138 | 0 | 53,920 | 371 | 428 |

| | | | | | | | |
|---|---|---|---|---|---|---|---|
| NegK-240404 | 14969 | 8027 | 2885 | 16542 | 78,954 | 3,010 | 3,174 |
| NegK-Extrakt-240705 | 255 | 255 | 760 | 463 | 24,960 | 784 | 815 |
| 15-sau6-3 | 0 | 0 | 0 | 1 | 343,227 | 61 | 102 |
| 4-eco6-1 | 0 | 0 | 0 | 1 | 133,250 | 4 | 15 |
| NegK-20230929 | 0 | 0 | 0 | 3 | 417,348 | 7 | 355 |
| NegK-20240216 | 0 | 0 | 0 | 0 | 160,880 | 85 | 166 |
| NegK-241108 | 0 | 0 | 0 | 8 | 84,994 | 17 | 22 |
| NegK-Extrakt-240412 | 0 | 0 | 0 | 0 | 38,323 | 4 | 20 |
| NegK-Extrakt-240517 | 0 | 0 | 0 | 0 | 17,700 | 0 | 24 |
| NegK-Extrakt-241101 | 0 | 0 | 0 | 45 | 159,504 | 38 | 65 |
| NegK-Extrakt-250115 | 0 | 0 | 0 | 3 | 278,302 | 5 | 31 |
| NFW-NegK-1 | 0 | 0 | 0 | 0 | 152,532 | 0 | 0 |
| NFW-NegK-3 | 0 | 0 | 0 | 0 | 12,387 | 0 | 0 |
| NFW-NegK-4 | 0 | 0 | 0 | 0 | 86,494 | 0 | 0 |
| NFW-NegK-2 | 0 | 0 | 0 | 0 | 24,544 | 0 | 0 |

**Supplementary Table 2**. Identified species in dilution series, their assigned read numbers, and BLAST-based species assignments of ONT raw reads.

| Sample_ID | species | Nbr. Reads Illumina | Nbr. Reads Illumina subsample | Nbr. Reads ONT assigned to consensus | Nbr. ONT raw reads |
|---|---|---|---|---|---|
| 1-eco5-1 | Actinomyces oris | 206 | 109 | 0 | 68 |
| 1-eco5-1 | Cutibacterium acnes | 378 | 228 | 0 | 100 |
| 1-eco5-1 | Escherichia sp. | 13834 | 8520 | 2928 | 3051 |
| 10-sau5-1 | Cutibacterium acnes | 447 | 448 | 0 | 16 |
| 10-sau5-1 | Staphylococcus aureus | 5053 | 5053 | 123 | 128 |
| 11-sau5-2 | Staphylococcus aureus | 4317 | 4317 | 223 | 229 |
| 12-sau5-3 | Staphylococcus aureus | 2776 | 2774 | 112 | 116 |
| 13-sau6-1 | Cutibacterium acnes | 241 | 241 | 0 | 33 |
| 13-sau6-1 | Staphylococcus aureus | 1447 | 1447 | 0 | 72 |
| 14-sau6-2 | Staphylococcus aureus | 3142 | 3141 | 0 | 66 |
| 16-sau7-1 | Cnuibacter physcomitrellae | 1090 | 1090 | 0 | 4 |
| 17-sau7-2 | Corynebacterium mucifaciens | 519 | 519 | 0 | 63 |
| 17-sau7-2 | Corynebacterium pseudokroppenstedtii | 416 | 416 | 0 | 82 |
| 18-sau7-3 | Corynebacterium afermentans | 830 | 830 | 0 | 14 |
| 19-mix5-1 | Escherichia sp. | 17391 | 5563 | 1793 | 1873 |
| 19-mix5-1 | Staphylococcus aureus | 9855 | 3042 | 438 | 454 |
| 2-eco5-2 | Cutibacterium acnes | 326 | 315 | 0 | 54 |
| 2-eco5-2 | Escherichia sp. | 8935 | 8456 | 1234 | 1487 |
| 20-mix5-2 | Escherichia sp. | 14858 | 5852 | 1309 | 1475 |
| 20-mix5-2 | Staphylococcus aureus | 7054 | 2797 | 299 | 306 |
| 21-mix5-3 | Escherichia sp. | 11953 | 7715 | 2989 | 3144 |
| 21-mix5-3 | Staphylococcus aureus | 1586 | 919 | 206 | 212 |
| 22-mix6-1 | Escherichia sp. | 8202 | 6112 | 2802 | 2928 |
| 22-mix6-1 | Staphylococcus aureus | 3524 | 2568 | 680 | 695 |
| 23-mix6-2 | Escherichia sp. | 11702 | 4890 | 2153 | 2269 |
| 23-mix6-2 | Staphylococcus aureus | 9346 | 3701 | 863 | 892 |
| 24-mix6-3 | Escherichia sp. | 14267 | 5555 | 1323 | 1474 |
| 24-mix6-3 | Staphylococcus aureus | 8135 | 3211 | 370 | 389 |
| 25-mix7-1 | Corynebacterium pseudokroppenstedtii | 113 | 103 | 0 | 9 |
| 25-mix7-1 | Escherichia sp. | 15359 | 6586 | 550 | 666 |
| 25-mix7-1 | Staphylococcus aureus | 2969 | 1246 | 0 | 61 |
| 25-mix7-1 | Streptococcus oralis | 1809 | 730 | 0 | 36 |
| 25-mix7-1 | Thermus parvatiensis | 154 | 0 | 0 | 8 |
| 26-mix7-2 | Escherichia sp. | 9654 | 7062 | 0 | 263 |
| 26-mix7-2 | Streptococcus parasanguinis | 2389 | 1759 | 0 | 37 |
| 27-mix7-3 | Escherichia sp. | 9556 | 7240 | 754 | 922 |
| 27-mix7-3 | Halomonas xinjiangensis | 214 | 108 | 0 | 20 |
| 27-mix7-3 | Staphylococcus aureus | 1769 | 1322 | 110 | 118 |

| | | | | | |
|---|---|---|---|---|---|
| 3-eco5-3 | Escherichia sp. | 9527 | 8912 | 822 | 993 |
| 5-eco6-2 | Corynebacterium kroppenstedtii | 2242 | 2046 | 0 | 44 |
| 5-eco6-2 | Escherichia sp. | 750 | 667 | 0 | 19 |
| 5-eco6-2 | Paracoccus hibiscisoli | 5520 | 4546 | 0 | 91 |
| 6-eco6-3 | Cutibacterium acnes | 165 | 165 | 0 | 42 |
| 7-eco7-1 | Cutibacterium acnes | 786 | 786 | 0 | 30 |
| 7-eco7-1 | Rhizobium daejeonense | 1406 | 1406 | 0 | 25 |
| 8-eco7-2 | Cutibacterium acnes | 142 | 142 | 0 | 35 |
| 9-eco7-3 | Escherichia sp. | 1050 | 1050 | 0 | 61 |
| 1-eco5-1 | Actinomyces naeslundii | NA | NA | NA | 2 |
| 1-eco5-1 | Cutibacterium avidum | NA | NA | NA | 2 |
| 1-eco5-1 | Enterobacteriaceae bacterium | NA | NA | NA | 10 |
| 1-eco5-1 | Erwinia psidii | NA | NA | NA | 59 |
| 1-eco5-1 | Pectobacterium carotovorum | NA | NA | NA | 11 |
| 10-sau5-1 | Staphylococcus haemolyticus | NA | NA | NA | 2 |
| 11-sau5-2 | Escherichia sp. | NA | NA | NA | 2 |
| 11-sau5-2 | Staphylococcus saccharolyticus | NA | NA | NA | 2 |
| 12-sau5-3 | Escherichia sp. | NA | NA | NA | 2 |
| 13-sau6-1 | Escherichia sp. | NA | NA | NA | 3 |
| 13-sau6-1 | Streptococcus agalactiae | NA | NA | NA | 2 |
| 15-sau6-3 | Escherichia sp. | NA | NA | NA | 42 |
| 15-sau6-3 | Staphylococcus aureus | NA | NA | NA | 10 |
| 15-sau6-3 | Streptococcus agalactiae | NA | NA | NA | 6 |
| 16-sau7-1 | Erwinia psidii | NA | NA | NA | 2 |
| 16-sau7-1 | Escherichia sp. | NA | NA | NA | 34 |
| 16-sau7-1 | Leifsonia aquatica | NA | NA | NA | 6 |
| 16-sau7-1 | Microbacterium arborescens | NA | NA | NA | 2 |
| 16-sau7-1 | Microbacterium paraoxydans | NA | NA | NA | 2 |
| 16-sau7-1 | Staphylococcus aureus | NA | NA | NA | 10 |
| 16-sau7-1 | Streptococcus agalactiae | NA | NA | NA | 6 |
| 17-sau7-2 | Corynebacterium afermentans | NA | NA | NA | 2 |
| 17-sau7-2 | Corynebacterium kroppenstedtii | NA | NA | NA | 6 |
| 17-sau7-2 | Escherichia sp. | NA | NA | NA | 38 |
| 17-sau7-2 | Staphylococcus aureus | NA | NA | NA | 7 |
| 17-sau7-2 | Streptococcus agalactiae | NA | NA | NA | 5 |
| 18-sau7-3 | Corynebacterium coyleae | NA | NA | NA | 4 |
| 18-sau7-3 | Corynebacterium mucifaciens | NA | NA | NA | 5 |
| 18-sau7-3 | Corynebacterium pilbarense | NA | NA | NA | 20 |
| 18-sau7-3 | Corynebacterium ureicelerivorans | NA | NA | NA | 18 |
| 18-sau7-3 | Escherichia sp. | NA | NA | NA | 28 |
| 18-sau7-3 | Halomonas xinjiangensis | NA | NA | NA | 2 |
| 18-sau7-3 | Staphylococcus aureus | NA | NA | NA | 9 |
| 18-sau7-3 | Streptococcus agalactiae | NA | NA | NA | 6 |
| 19-mix5-1 | Enterobacteriaceae bacterium | NA | NA | NA | 8 |
| 19-mix5-1 | Erwinia psidii | NA | NA | NA | 43 |
| 19-mix5-1 | Pectobacterium carotovorum | NA | NA | NA | 3 |

| | | | | | |
|---|---|---|---|---|---|
| 19-mix5-1 | Staphylococcus carnosus | NA | NA | NA | 5 |
| 19-mix5-1 | Staphylococcus haemolyticus | NA | NA | NA | 5 |
| 19-mix5-1 | Staphylococcus hominis | NA | NA | NA | 3 |
| 19-mix5-1 | Staphylococcus warneri | NA | NA | NA | 2 |
| 19-mix5-1 | Streptococcus agalactiae | NA | NA | NA | 3 |
| 19-mix5-1 | Streptococcus parasanguinis | NA | NA | NA | 3 |
| 2-eco5-2 | Enterobacteriaceae bacterium | NA | NA | NA | 2 |
| 2-eco5-2 | Erwinia psidii | NA | NA | NA | 29 |
| 2-eco5-2 | Pectobacterium carotovorum | NA | NA | NA | 5 |
| 20-mix5-2 | Enterobacteriaceae bacterium | NA | NA | NA | 5 |
| 20-mix5-2 | Erwinia psidii | NA | NA | NA | 37 |
| 20-mix5-2 | Staphylococcus haemolyticus | NA | NA | NA | 2 |
| 20-mix5-2 | Streptococcus agalactiae | NA | NA | NA | 4 |
| 21-mix5-3 | Brevibacterium paucivorans | NA | NA | NA | 11 |
| 21-mix5-3 | Cutibacterium acnes | NA | NA | NA | 18 |
| 21-mix5-3 | Enterobacteriaceae bacterium | NA | NA | NA | 7 |
| 21-mix5-3 | Erwinia psidii | NA | NA | NA | 71 |
| 21-mix5-3 | Pectobacterium carotovorum | NA | NA | NA | 13 |
| 21-mix5-3 | Staphylococcus haemolyticus | NA | NA | NA | 2 |
| 21-mix5-3 | Staphylococcus simiae | NA | NA | NA | 3 |
| 21-mix5-3 | Streptococcus agalactiae | NA | NA | NA | 13 |
| 22-mix6-1 | Cutibacterium acnes | NA | NA | NA | 14 |
| 22-mix6-1 | Enterobacteriaceae bacterium | NA | NA | NA | 2 |
| 22-mix6-1 | Erwinia psidii | NA | NA | NA | 72 |
| 22-mix6-1 | Pectobacterium carotovorum | NA | NA | NA | 9 |
| 22-mix6-1 | Staphylococcus carnosus | NA | NA | NA | 5 |
| 22-mix6-1 | Staphylococcus haemolyticus | NA | NA | NA | 2 |
| 22-mix6-1 | Staphylococcus warneri | NA | NA | NA | 3 |
| 22-mix6-1 | Streptococcus agalactiae | NA | NA | NA | 3 |
| 23-mix6-2 | Caminibacter mediatlanticus | NA | NA | NA | 2 |
| 23-mix6-2 | Cutibacterium acnes | NA | NA | NA | 4 |
| 23-mix6-2 | Enterobacteriaceae bacterium | NA | NA | NA | 8 |
| 23-mix6-2 | Erwinia psidii | NA | NA | NA | 49 |
| 23-mix6-2 | Pectobacterium carotovorum | NA | NA | NA | 8 |
| 23-mix6-2 | Staphylococcus caprae | NA | NA | NA | 2 |
| 23-mix6-2 | Staphylococcus carnosus | NA | NA | NA | 5 |
| 23-mix6-2 | Staphylococcus haemolyticus | NA | NA | NA | 5 |
| 23-mix6-2 | Staphylococcus hominis | NA | NA | NA | 2 |
| 23-mix6-2 | Streptococcus agalactiae | NA | NA | NA | 4 |
| 24-mix6-3 | Caminibacter mediatlanticus | NA | NA | NA | 2 |
| 24-mix6-3 | Cutibacterium acnes | NA | NA | NA | 2 |
| 24-mix6-3 | Enterobacteriaceae bacterium | NA | NA | NA | 4 |
| 24-mix6-3 | Erwinia psidii | NA | NA | NA | 26 |
| 24-mix6-3 | Pectobacterium carotovorum | NA | NA | NA | 4 |
| 24-mix6-3 | Staphylococcus caprae | NA | NA | NA | 2 |
| 24-mix6-3 | Staphylococcus warneri | NA | NA | NA | 2 |

| | | | | | |
|---|---|---|---|---|---|
| 24-mix6-3 | Streptococcus agalactiae | NA | NA | NA | 4 |
| 25-mix7-1 | Enterobacteriaceae bacterium | NA | NA | NA | 2 |
| 25-mix7-1 | Erwinia psidii | NA | NA | NA | 12 |
| 25-mix7-1 | Streptococcus agalactiae | NA | NA | NA | 8 |
| 25-mix7-1 | Streptococcus humanilactis | NA | NA | NA | 2 |
| 25-mix7-1 | Streptococcus mitis | NA | NA | NA | 6 |
| 26-mix7-2 | Enterobacteriaceae bacterium | NA | NA | NA | 2 |
| 26-mix7-2 | Erwinia psidii | NA | NA | NA | 5 |
| 26-mix7-2 | Staphylococcus aureus | NA | NA | NA | 6 |
| 26-mix7-2 | Streptococcus agalactiae | NA | NA | NA | 9 |
| 27-mix7-3 | Erwinia psidii | NA | NA | NA | 26 |
| 27-mix7-3 | Halomonas saudii | NA | NA | NA | 2 |
| 27-mix7-3 | Streptococcus agalactiae | NA | NA | NA | 5 |
| 3-eco5-3 | Enterobacteriaceae bacterium | NA | NA | NA | 3 |
| 3-eco5-3 | Erwinia psidii | NA | NA | NA | 14 |
| 3-eco5-3 | Pectobacterium carotovorum | NA | NA | NA | 2 |
| 4-eco6-1 | Staphylococcus aureus | NA | NA | NA | 2 |
| 4-eco6-1 | Streptococcus agalactiae | NA | NA | NA | 2 |
| 5-eco6-2 | Paracoccus halotolerans | NA | NA | NA | 2 |
| 5-eco6-2 | Paracoccus liaowanqingii | NA | NA | NA | 3 |
| 8-eco7-2 | Escherichia sp. | NA | NA | NA | 5 |
| 9-eco7-3 | Halomonas xinjiangensis | NA | NA | NA | 5 |
| 9-eco7-3 | Propionibacterium westphaliense | NA | NA | NA | 18 |

**Supplementary Table 3.** Clinical samples, clinical evaluation, antibiotic treatment, culture results and sample material.

| Sample_ID | Antibiotic treatment before sampling | clinical evaluation of sequencing | clinical information | sample type | culture results | agreement culture to sequencing | positive culture results from different specimens of same patient |
|---|---|---|---|---|---|---|---|
| I_eubac_clinic_001 | Unknown | contamination | check before implantation of a LVAD | blood, venous | unknown | | |
| I_eubac_clinic_002 | No | contamination | Reative arthtitis | puncture, joint, knee, right | negative | | |
| I_eubac_clinic_003 | Yes | reasonable | Prosthetic mitral valve endocarditis | biopsy, aortic valve | negative | | |
| I_eubac_clinic_004 | Yes | contamination | pleural sarcoma | biopsy, parietal pleura, right | negative | | |
| I_eubac_clinic_005 | Yes | reasonable | Osteosynthesis (shoulder) infection | biopsy, shoulder, left | negative | | |
| I_eubac_clinic_006 | Yes | reasonable | Osteosynthesis (shoulder) infection | biopsy, shoulder, left | negative | | |
| I_eubac_clinic_007 | No | reasonable | Mycotic aneurysm | biopsy, aorta | negative | | available |
| I_eubac_clinic_008 | Yes | reasonable | Prostehtic joint infetion | biopsy, femoral, left | negative | | |
| I_eubac_clinic_009 | Yes | reasonable | Prostehtic joint infetion | biopsy, femoral, left | negative | | |
| I_eubac_clinic_010 | Yes | reasonable | Prostehtic joint infetion | biopsy, femoral, left | negative | | |
| I_eubac_clinic_011 | Yes | reasonable | Prostehtic joint infetion | biopsy, femoral, left | negative | | |
| I_eubac_clinic_012 | Yes | reasonable | Prostehtic joint infetion | biopsy, femoral, left | negative | | |
| I_eubac_clinic_013 | Yes | reasonable | Prostehtic joint infetion | biopsy, joint, hip, left | negative | | |
| I_eubac_clinic_014 | Yes | reasonable | Prostehtic joint infetion | biopsy, joint, hip, left | negative | | |
| I_eubac_clinic_015 | Yes | reasonable | Prostehtic joint infetion | biopsy, joint, hip, left | negative | | |
| I_eubac_clinic_016 | Yes | reasonable | Prostehtic joint infetion | biopsy, joint, hip, left | negative | | |
| I_eubac_clinic_017 | No | reasonable | Aortic graft infection | blood, venous | unknown | | |
| I_eubac_clinic_018 | Yes | reasonable | pneumonia | puncture, thorax, left | negative | | |
| I_eubac_clinic_019 | No | contamination | arthritis knee (autoinflammation, seronegative spondylarthtis) | biopsy, joint, hip, left | negative | | |
| I_eubac_clinic_020 | No | reasonable | Spondylodesis | biopsy, spine, lumbar | negative | | |
| I_eubac_clinic_021 | No | reasonable | Spondylodesis | biopsy, spine, lumbar | negative | | |
| I_eubac_clinic_022 | No | reasonable | Spondylodesis | biopsy, spine, lumbar | negative | | |
| I_eubac_clinic_023 | Yes | reasonable | Spondylodesis | puncture, pleura | positive | yes | |
| I_eubac_clinic_024 | Yes | reasonable | Pneumonia | puncture, pleura left | negative | | |
| I_eubac_clinic_025 | No | contamination | Biopsy calcaneus. Clinically and histopathology no osteomyelitis. | biopsy, heel, left | negative | | |
| I_eubac_clinic_026 | Yes | reasonable | VP shunt infeciton | cerebrospinal fluid, brain | negative | | available |
| I_eubac_clinic_027 | Yes | reasonable | VP shunt infeciton | cerebrospinal fluid, lumbar puncture | negative | | available |
| I_eubac_clinic_028 | Yes | reasonable | Spondylodesis | biopsy, spine, lumbar | negative | | |

| | | | | | | | |
|---|---|---|---|---|---|---|---|
| I_eubac_clinic_029 | Unknown | reasonable | Prosthetic joint infection (knee) | puncture, joint, knee, left | negative | | |
| I_eubac_clinic_030 | Unknown | reasonable | Prosthetic joint infection (knee) | biopsy, joint, knee, left | negative | | |
| I_eubac_clinic_031 | Unknown | reasonable | Prosthetic joint infection (knee) | biopsy, joint, knee, left | negative | | |
| I_eubac_clinic_032 | Unknown | reasonable | Prosthetic joint infection (knee) | biopsy, joint, elbow, right | negative | | |
| I_eubac_clinic_033 | Unknown | reasonable | Prosthetic joint infection (knee) | biopsy, joint, elbow, right | negative | | |
| I_eubac_clinic_034 | Yes | reasonable | Spondylodiscitis | biopsy, lumbar vertebra | negative | | |
| I_eubac_clinic_035 | Yes | reasonable | Spondylodiscitis | biopsy, lumbar vertebra | negative | | |
| I_eubac_clinic_036 | Yes | reasonable | Spondylodiscitis | biopsy, lumbar vertebra | negative | | |
| I_eubac_clinic_037 | Yes | reasonable | Osteosynthesis ass infection | biopsy, knee, left | negative | | |
| I_eubac_clinic_038 | Yes | reasonable | Osteosynthesis ass infection | biopsy, joint, knee, right | negative | | |
| I_eubac_clinic_039 | No | contamination | langerhans histiocytosis | blood, venous | unknown | | |
| I_eubac_clinic_040 | Yes | reasonable | Prosthetic joint infection (knee) | puncture, joint, knee, right | negative | | |
| I_eubac_clinic_041 | Yes | reasonable | Mastoiditis | puncture, cerebrospinal fluid, lumbar puncture | negative | | |
| I_eubac_clinic_042 | Yes | reasonable | Spondylodiscitis | biopsy, lumbar vertebra | positive | yes | |
| I_eubac_clinic_043 | Yes | reasonable | Spondylodiscitis | biopsy, lumbar vertebra | positive | yes | |
| I_eubac_clinic_044 | Yes | reasonable | Spondylodiscitis | biopsy, lumbar vertebra | positive | yes | |
| I_eubac_clinic_045 | No | reasonable | Subdural empyema | biopsy, head | negative | | |
| I_eubac_clinic_046 | Yes | contamination | Spondylodiscitis | biopsy, vertebra of the chest | negative | | |
| I_eubac_clinic_047 | Yes | contamination | Spondylodiscitis | biopsy, vertebra of the chest | negative | | |
| I_eubac_clinic_048 | Yes | reasonable | Spondylodiscitis | biopsy, vertebra of the chest | negative | | |
| I_eubac_clinic_049 | Yes | reasonable | Spondylodiscitis | biopsy, vertebra of the chest | negative | | |
| I_eubac_clinic_050 | No | reasonable | Osteosynthesis ass infection pelvis | puncture, pelvis region | negative | | |
| I_eubac_clinic_051 | No | contamination | Juvenile arthritis | puncture, knee, right | negative | | |
| I_eubac_clinic_052 | No | reasonable | Septic gonarathrtis | puncture, joint, knee, right | negative | | |
| I_eubac_clinic_053 | Yes | reasonable | Prosthetic joint infection (hip) | biopsy, hip, left | negative | | |
| I_eubac_clinic_054 | No | reasonable | Liver cirrhosis | blood, venous | positive | yes | |
| I_eubac_clinic_055 | No | reasonable | Prosthetic joint infection (hip) | biopsy, joint, hip, right | negative | | |
| I_eubac_clinic_056 | No | reasonable | Prosthetic joint infection (hip) | biopsy, joint, hip, right | negative | | |
| I_eubac_clinic_057 | Yes | contamination | NA | blood, venous | positive | no | |
| I_eubac_clinic_058 | Yes | reasonable | Bursitis | biopsy, joint capsule | negative | | |
| I_eubac_clinic_059 | Yes | reasonable | Pleuraempyem | biopsy, pleura | negative | | |
| I_eubac_clinic_060 | Yes | contamination | Spondylodesis | biopsy, spine, lumbar | positive | no | |
| I_eubac_clinic_061 | Yes | contamination | Spondylodesis | biopsy, spine, lumbar | positive | no | |
| I_eubac_clinic_062 | Yes | contamination | Spondylodesis | biopsy, spine, lumbar | positive | no | |
| I_eubac_clinic_063 | Yes | reasonable | Osteomyelitis mandibula | biopsy, mandible, right | negative | | available |
| I_eubac_clinic_064 | Yes | reasonable | Prosthetic joint infection (knee) | biopsy, joint, knee, left | negative | | |

| | | | | | | | |
|---|---|---|---|---|---|---|---|
| I_eubac_clinic_065 | Yes | reasonable | Prosthetic joint infection (knee) | biopsy, joint, knee, left | negative | | |
| I_eubac_clinic_066 | Yes | reasonable | Prosthetic joint infection (knee) | biopsy, joint, knee, left | negative | | |
| I_eubac_clinic_067 | Yes | contamination | Spondylodesis | biopsy, vertebra, cervical | negative | | |
| I_eubac_clinic_068 | Yes | contamination | Spondylodesis | biopsy, vertebra, cervical | negative | | |
| I_eubac_clinic_069 | Yes | contamination | Prosthetic valve endocarditis | biopsy, popliteal artery, left | negative | | |
| I_eubac_clinic_070 | Yes | reasonable | Prosthetic joint infection (hip) | biopsy, joint, hip, left | negative | | |
| I_eubac_clinic_071 | Yes | contamination | Prosthetic joint infection (hip) | biopsy, joint, hip, left | negative | | |
| I_eubac_clinic_072 | Yes | reasonable | Prosthetic joint infection (hip) | biopsy, joint, hip, left | negative | | |
| I_eubac_clinic_073 | Yes | reasonable | Prosthetic valve endocarditis | biopsy, aortic valve | positive | yes | |
| I_eubac_clinic_074 | Yes | reasonable | Prosthetic joint infection (knee) | biopsy, joint, shoulder, right | negative | | |
| I_eubac_clinic_075 | Yes | reasonable | Prosthetic joint infection (hip) | biopsy, hip joint, left | negative | | |
| I_eubac_clinic_076 | Yes | reasonable | Prosthetic joint infection (hip) | biopsy, hip joint, left | negative | | |
| I_eubac_clinic_077 | Yes | reasonable | Prosthetic joint infection (hip) | biopsy, hip joint, left | negative | | |
| I_eubac_clinic_078 | Yes | reasonable | retroperitoneal lymphadenitis | biopsy, peritoneum, pelvis | negative | | |
| I_eubac_clinic_079 | Yes | reasonable | Prosthetic valve endocarditis | biopsy, mitral valve | negative | | |
| I_eubac_clinic_080 | Yes | reasonable | Mycotic aneurysm | biopsy, aorta | negative | | |
| I_eubac_clinic_081 | Yes | reasonable | Native valve endocarditis | biopsy, mitral valve | negative | | available |
| I_eubac_clinic_082 | Yes | reasonable | biopsy of shoulder. Diss Gr. B streptococci infection | puncture, joint, shoulder, left | negative | | available |
| I_eubac_clinic_083 | Yes | reasonable | Osteosynthesis ass infection ankle. | biopsy, leg, shank, left | negative | | |
| I_eubac_clinic_084 | Yes | reasonable | Osteosynthesis ass infection ankle. | biopsy, leg, shank, left | negative | | |
| I_eubac_clinic_085 | No | reasonable | Osteosynthesis ass infection humerus | biopsy, joint, shoulder, left | positive | yes | |
| I_eubac_clinic_086 | No | reasonable | Prosthetic joint infection (hip) | puncture, joint, hip, left | positive | yes | |
| I_eubac_clinic_087 | No | contamination | knee biopsies | biopsy: subcutaneous collection knee right | negative | | |
| I_eubac_clinic_088 | No | contamination | knee prothesis | bone biopsy: ossifications knee right | negative | | |
| I_eubac_clinic_089 | No | contamination | knee prothesis | biopsy synovia: posterior capsule knee right | negative | | |
| I_eubac_clinic_090 | Yes | reasonable | NA | biopsy bursa synovialis: synovia knee right | negative | | |
| I_eubac_clinic_091 | Yes | reasonable | NA | biopsy: synovia knee right | negative | | |
| I_eubac_clinic_092 | No | reasonable | knee prothesis | puncture bursa synovialis: knee right | negative | | |
| I_eubac_clinic_093 | No | contamination | knee prothesis | biopsy: intercondylar notch knee right | negative | | |
| I_eubac_clinic_094 | No | contamination | knee prothesis | synovial fluid: knee left | negative | | |
| I_eubac_clinic_095 | Yes | contamination | Osteosynthesis infection tibia | biopsy: wound deep tibia right | negative | | available |
| I_eubac_clinic_096 | No | contamination | spondylodiscitis | bone biopsy: spinal disc L3/4 | negative | | |

| | | | | | |
|---|---|---|---|---|---|
| I_eubac_clinic_097 | No | reasonable | Palacos infection | biopsy wound deep: neodura | negative |
| I_eubac_clinic_098 | Yes | reasonable | hip infection | bone biopsy: femoral neck hip left | negative |
| I_eubac_clinic_099 | Yes | reasonable | pneumonia | pleural liquid: pleural empyema right | negative |
| I_eubac_clinic_100 | Yes | reasonable | iliac crest abscess | biopsy wound deep | negative |
| I_eubac_clinic_101 | Yes | reasonable | septic omarthritis | biopsy: subacromial, shoulder left | negative |

**Supplementary Table 4.** Species detected in clinical samples and their assigned read numbers for the respective sequencing methods.

| species | Nbr. Reads Illumina | Nbr. Reads Illumina subsample | Nbr. Reads ONT | Sample_ID |
|---|---|---|---|---|
| Corynebacterium kroppenstedtii | 0 | 0 | 332 | I_eubac_clinic_001 |
| Corynebacterium pseudokroppenstedtii | 18999 | 3905 | 2162 | I_eubac_clinic_001 |
| Cutibacterium acnes | 17205 | 3422 | 2437 | I_eubac_clinic_001 |
| Cutibacterium granulosum | 103 | 0 | 0 | I_eubac_clinic_001 |
| Salibacter halophilus | 4068 | 902 | 295 | I_eubac_clinic_001 |
| Cutibacterium acnes | 26430 | 5117 | 4660 | I_eubac_clinic_002 |
| Lawsonella clevelandensis | 10393 | 1911 | 1378 | I_eubac_clinic_002 |
| Micrococcus luteus | 6149 | 1221 | 911 | I_eubac_clinic_002 |
| Corynebacterium kroppenstedtii | 0 | 0 | 149 | I_eubac_clinic_003 |
| Corynebacterium pseudokroppenstedtii | 5271 | 2132 | 937 | I_eubac_clinic_003 |
| Cutibacterium acnes | 8785 | 3637 | 1896 | I_eubac_clinic_003 |
| Lawsonella clevelandensis | 6023 | 2520 | 959 | I_eubac_clinic_003 |
| Actinomyces viscosus | 1135 | 175 | 459 | I_eubac_clinic_004 |
| Corynebacterium kroppenstedtii | 2230 | 390 | 768 | I_eubac_clinic_004 |
| Cutibacterium acnes | 9132 | 1540 | 3709 | I_eubac_clinic_004 |
| Ezakiella coagulans | 998 | 169 | 224 | I_eubac_clinic_004 |
| Lawsonella clevelandensis | 1837 | 314 | 555 | I_eubac_clinic_004 |
| Rothia dentocariosa | 3019 | 625 | 1723 | I_eubac_clinic_004 |
| Streptococcus oralis | 3179 | 559 | 730 | I_eubac_clinic_004 |
| Bacillus mycoides | 708 | 123 | 0 | I_eubac_clinic_005 |
| Cutibacterium acnes | 4995 | 970 | 1094 | I_eubac_clinic_005 |
| Cutibacterium granulosum | 17006 | 4931 | 7458 | I_eubac_clinic_005 |
| Cutibacterium namnetense | 0 | 0 | 171 | I_eubac_clinic_005 |
| Lactococcus lactis | 583 | 0 | 0 | I_eubac_clinic_005 |
| Leptothrix mobilis | 540 | 0 | 0 | I_eubac_clinic_005 |
| Corynebacterium kroppenstedtii | 1441 | 323 | 236 | I_eubac_clinic_006 |
| Corynebacterium pseudokroppenstedtii | 224 | 0 | 0 | I_eubac_clinic_006 |
| Corynebacterium striatum | 546 | 0 | 104 | I_eubac_clinic_006 |
| Cutibacterium acnes | 4433 | 894 | 998 | I_eubac_clinic_006 |
| Cutibacterium granulosum | 16340 | 3193 | 7286 | I_eubac_clinic_006 |
| Cutibacterium namnetense | 0 | 0 | 198 | I_eubac_clinic_006 |
| Streptococcus pneumoniae | 584 | 0 | 0 | I_eubac_clinic_007 |
| Streptococcus pseudopneumoniae | 58173 | 8512 | 8633 | I_eubac_clinic_007 |
| Streptococcus dysgalactiae | 45174 | 8738 | 8908 | I_eubac_clinic_008 |
| Streptococcus dysgalactiae | 37745 | 8752 | 7854 | I_eubac_clinic_009 |
| Streptococcus dysgalactiae | 26070 | 8809 | 8913 | I_eubac_clinic_010 |
| Streptococcus dysgalactiae | 46282 | 8722 | 8997 | I_eubac_clinic_011 |
| Streptococcus dysgalactiae | 40639 | 8775 | 8996 | I_eubac_clinic_012 |
| Cutibacterium acnes | 560 | 119 | 0 | I_eubac_clinic_013 |
| Streptococcus dysgalactiae | 46740 | 8738 | 6902 | I_eubac_clinic_013 |
| Cutibacterium acnes | 114 | 0 | 0 | I_eubac_clinic_014 |
| Streptococcus dysgalactiae | 47418 | 8799 | 8887 | I_eubac_clinic_014 |

| | | | | |
|---|---|---|---|---|
| Streptococcus dysgalactiae | 28972 | 8756 | 2090 | I_eubac_clinic_015 |
| Streptococcus dysgalactiae | 44726 | 8833 | 8903 | I_eubac_clinic_016 |
| Staphylococcus caprae | 54765 | 8188 | 5705 | I_eubac_clinic_017 |
| Staphylococcus epidermidis | 3898 | 598 | 535 | I_eubac_clinic_017 |
| Streptococcus pneumoniae | 631 | 167 | 0 | I_eubac_clinic_018 |
| Streptococcus pseudopneumoniae | 31393 | 8481 | 4292 | I_eubac_clinic_018 |
| Corynebacterium segmentosum | 3432 | 1161 | 119 | I_eubac_clinic_019 |
| Cutibacterium acnes | 3992 | 1331 | 281 | I_eubac_clinic_019 |
| Lawsonella clevelandensis | 4467 | 1586 | 254 | I_eubac_clinic_019 |
| Moraxella osloensis | 2618 | 881 | 132 | I_eubac_clinic_019 |
| Oceanicella actignis | 7472 | 2513 | 341 | I_eubac_clinic_019 |
| Staphylococcus caprae | 2570 | 896 | 0 | I_eubac_clinic_019 |
| Cutibacterium acnes | 39782 | 6763 | 8683 | I_eubac_clinic_020 |
| Cutibacterium namnetense | 0 | 0 | 116 | I_eubac_clinic_020 |
| Cutibacterium acnes | 32141 | 8165 | 8774 | I_eubac_clinic_021 |
| Cutibacterium namnetense | 0 | 0 | 110 | I_eubac_clinic_021 |
| Cutibacterium acnes | 32223 | 7853 | 8865 | I_eubac_clinic_022 |
| Cutibacterium namnetense | 0 | 0 | 108 | I_eubac_clinic_022 |
| Staphylococcus caprae | 45237 | 7237 | 7266 | I_eubac_clinic_023 |
| Staphylococcus epidermidis | 7657 | 1331 | 1472 | I_eubac_clinic_023 |
| Streptococcus pneumoniae | 835 | 144 | 0 | I_eubac_clinic_024 |
| Streptococcus pseudopneumoniae | 49828 | 8719 | 8749 | I_eubac_clinic_024 |
| Dermacoccus nishinomiyaensis | 353 | 354 | 1395 | I_eubac_clinic_025 |
| Staphylococcus capitis | 862 | 862 | 1887 | I_eubac_clinic_025 |
| Staphylococcus warneri | 1148 | 1148 | 3307 | I_eubac_clinic_025 |
| Cutibacterium acnes | 434 | 400 | 334 | I_eubac_clinic_026 |
| Erwinia psidii | 0 | 0 | 180 | I_eubac_clinic_026 |
| Escherichia sp. | 8931 | 8136 | 6110 | I_eubac_clinic_026 |
| Pseudonocardia alni | 256 | 226 | 208 | I_eubac_clinic_026 |
| Corynebacterium kroppenstedtii | 262 | 263 | 189 | I_eubac_clinic_027 |
| Cutibacterium acnes | 187 | 187 | 115 | I_eubac_clinic_027 |
| Escherichia sp. | 1735 | 1736 | 882 | I_eubac_clinic_027 |
| Lawsonella clevelandensis | 1469 | 1469 | 782 | I_eubac_clinic_027 |
| Sphingomonas carotinifaciens | 751 | 751 | 303 | I_eubac_clinic_027 |
| Cnuibacter physcomitrellae | 766 | 220 | 0 | I_eubac_clinic_028 |
| Corynebacterium aurimucosum | 7845 | 2217 | 1018 | I_eubac_clinic_028 |
| Cutibacterium acnes | 16650 | 4623 | 2490 | I_eubac_clinic_028 |
| Cutibacterium granulosum | 4911 | 1275 | 962 | I_eubac_clinic_028 |
| Cutibacterium namnetense | 0 | 0 | 135 | I_eubac_clinic_028 |
| Streptococcus australis | 0 | 0 | 114 | I_eubac_clinic_029 |
| Streptococcus humanilactis | 0 | 0 | 197 | I_eubac_clinic_029 |
| Streptococcus infantis | 0 | 0 | 198 | I_eubac_clinic_029 |
| Streptococcus mitis | 0 | 0 | 472 | I_eubac_clinic_029 |
| Streptococcus oralis | 49629 | 8833 | 7406 | I_eubac_clinic_029 |
| Streptococcus toyakuensis | 0 | 0 | 328 | I_eubac_clinic_029 |
| Cutibacterium acnes | 179 | 0 | 0 | I_eubac_clinic_030 |

| | | | | |
|---|---|---|---|---|
| Streptococcus australis | 0 | 0 | 115 | I_eubac_clinic_030 |
| Streptococcus humanilactis | 0 | 0 | 189 | I_eubac_clinic_030 |
| Streptococcus infantis | 0 | 0 | 177 | I_eubac_clinic_030 |
| Streptococcus mitis | 0 | 0 | 491 | I_eubac_clinic_030 |
| Streptococcus oralis | 44169 | 8795 | 7302 | I_eubac_clinic_030 |
| Streptococcus toyakuensis | 0 | 0 | 411 | I_eubac_clinic_030 |
| Streptococcus australis | 0 | 0 | 131 | I_eubac_clinic_031 |
| Streptococcus humanilactis | 0 | 0 | 160 | I_eubac_clinic_031 |
| Streptococcus infantis | 0 | 0 | 137 | I_eubac_clinic_031 |
| Streptococcus mitis | 0 | 0 | 380 | I_eubac_clinic_031 |
| Streptococcus oralis | 41495 | 8891 | 6047 | I_eubac_clinic_031 |
| Streptococcus toyakuensis | 0 | 0 | 304 | I_eubac_clinic_031 |
| Streptococcus australis | 0 | 0 | 113 | I_eubac_clinic_032 |
| Streptococcus humanilactis | 0 | 0 | 180 | I_eubac_clinic_032 |
| Streptococcus infantis | 0 | 0 | 140 | I_eubac_clinic_032 |
| Streptococcus mitis | 0 | 0 | 363 | I_eubac_clinic_032 |
| Streptococcus oralis | 38912 | 8896 | 4561 | I_eubac_clinic_032 |
| Streptococcus toyakuensis | 0 | 0 | 306 | I_eubac_clinic_032 |
| Granulicatella elegans | 221 | 0 | 0 | I_eubac_clinic_033 |
| Streptococcus australis | 0 | 0 | 178 | I_eubac_clinic_033 |
| Streptococcus humanilactis | 0 | 0 | 244 | I_eubac_clinic_033 |
| Streptococcus infantis | 0 | 0 | 208 | I_eubac_clinic_033 |
| Streptococcus mitis | 0 | 0 | 535 | I_eubac_clinic_033 |
| Streptococcus oralis | 48755 | 8675 | 6881 | I_eubac_clinic_033 |
| Streptococcus toyakuensis | 0 | 0 | 508 | I_eubac_clinic_033 |
| Actinomycetales bacterium | 1208 | 256 | 270 | I_eubac_clinic_034 |
| Cutibacterium acnes | 504 | 107 | 0 | I_eubac_clinic_034 |
| Halopseudomonas formosensis | 921 | 194 | 129 | I_eubac_clinic_034 |
| Sphingomonas hengshuiensis | 7818 | 1622 | 889 | I_eubac_clinic_034 |
| Staphylococcus hominis | 2813 | 554 | 333 | I_eubac_clinic_034 |
| Streptococcus pyogenes | 29471 | 6005 | 4839 | I_eubac_clinic_034 |
| Streptococcus pyogenes | 32086 | 8865 | 5012 | I_eubac_clinic_035 |
| Sphingomonas hengshuiensis | 509 | 0 | 0 | I_eubac_clinic_036 |
| Streptococcus pyogenes | 44715 | 8718 | 8951 | I_eubac_clinic_036 |
| Staphylococcus cohnii | 0 | 0 | 216 | I_eubac_clinic_037 |
| Staphylococcus nepalensis | 0 | 0 | 362 | I_eubac_clinic_037 |
| Staphylococcus ureilyticus | 31767 | 8912 | 8267 | I_eubac_clinic_037 |
| Enterobacter hormaechei | 913 | 913 | 0 | I_eubac_clinic_038 |
| Klebsiella grimontii | 7336 | 7338 | 1056 | I_eubac_clinic_038 |
| Amniculibacterium aquaticum | 1089 | 347 | 0 | I_eubac_clinic_039 |
| Caldibacillus hisashii | 928 | 308 | 178 | I_eubac_clinic_039 |
| Cloacibacterium normanense | 1090 | 333 | 136 | I_eubac_clinic_039 |
| Corynebacterium vitaeruminis | 520 | 180 | 0 | I_eubac_clinic_039 |
| Enterococcus cecorum | 19790 | 6210 | 3415 | I_eubac_clinic_039 |
| Georgenia deserti | 0 | 0 | 152 | I_eubac_clinic_039 |
| Georgenia halophila | 841 | 158 | 0 | I_eubac_clinic_039 |

| | | | | |
|---|---|---|---|---|
| Microbacterium sediminis | 1288 | 396 | 240 | I_eubac_clinic_039 |
| Micrococcus antarcticus | 338 | 0 | 0 | I_eubac_clinic_039 |
| Staphylococcus hominis | 1042 | 363 | 144 | I_eubac_clinic_039 |
| Tepidimonas taiwanensis | 483 | 0 | 121 | I_eubac_clinic_039 |
| Staphylococcus lugdunensis | 24587 | 8828 | 1257 | I_eubac_clinic_040 |
| Streptococcus pneumoniae | 339 | 0 | 0 | I_eubac_clinic_041 |
| Streptococcus pseudopneumoniae | 43251 | 8766 | 8677 | I_eubac_clinic_041 |
| Streptococcus agalactiae | 22686 | 8875 | 4308 | I_eubac_clinic_042 |
| Streptococcus agalactiae | 27716 | 8896 | 8020 | I_eubac_clinic_043 |
| Streptococcus agalactiae | 24622 | 8885 | 6819 | I_eubac_clinic_044 |
| Fusobacterium naviforme | 8715 | 1630 | 1042 | I_eubac_clinic_045 |
| Fusobacterium nucleatum | 38983 | 5807 | 7659 | I_eubac_clinic_045 |
| Staphylococcus caprae | 14651 | 3759 | 364 | I_eubac_clinic_046 |
| Staphylococcus warneri | 7304 | 996 | 372 | I_eubac_clinic_046 |
| Corynebacterium terpenotabidum | 875 | 225 | 157 | I_eubac_clinic_047 |
| Corynebacterium variabile | 734 | 0 | 136 | I_eubac_clinic_047 |
| Cutibacterium acnes | 522 | 0 | 101 | I_eubac_clinic_047 |
| Erythrobacter cryptus | 683 | 118 | 116 | I_eubac_clinic_047 |
| Lachnoanaerobaculum gingivalis | 1824 | 329 | 299 | I_eubac_clinic_047 |
| Lawsonella clevelandensis | 1202 | 224 | 208 | I_eubac_clinic_047 |
| Phenylobacterium deserti | 520 | 118 | 0 | I_eubac_clinic_047 |
| Schlegelella aquatica | 2360 | 387 | 519 | I_eubac_clinic_047 |
| Staphylococcus aureus | 862 | 0 | 150 | I_eubac_clinic_047 |
| Staphylococcus caprae | 22776 | 4273 | 4314 | I_eubac_clinic_047 |
| Staphylococcus epidermidis | 11993 | 2264 | 2215 | I_eubac_clinic_047 |
| Streptococcus australis | 285 | 0 | 0 | I_eubac_clinic_047 |
| Streptococcus koreensis | 1139 | 271 | 185 | I_eubac_clinic_047 |
| Streptococcus oralis | 346 | 0 | 0 | I_eubac_clinic_047 |
| Micrococcus antarcticus | 1193 | 236 | 213 | I_eubac_clinic_048 |
| Staphylococcus aureus | 41653 | 8400 | 6674 | I_eubac_clinic_048 |
| Staphylococcus aureus | 42924 | 8822 | 1844 | I_eubac_clinic_049 |
| Enterococcus faecium | 21153 | 8709 | 7623 | I_eubac_clinic_050 |
| Enterococcus gallinarum | 0 | 0 | 595 | I_eubac_clinic_050 |
| Enterococcus rivorum | 0 | 0 | 181 | I_eubac_clinic_050 |
| Cutibacterium acnes | 6987 | 6987 | 208 | I_eubac_clinic_051 |
| Cutibacterium acnes | 13579 | 8549 | 207 | I_eubac_clinic_052 |
| Paracoccus hibiscisoli | 8453 | 3372 | 2248 | I_eubac_clinic_053 |
| Paracoccus liaowanqingii | 0 | 0 | 130 | I_eubac_clinic_053 |
| Staphylococcus aureus | 15930 | 4357 | 2485 | I_eubac_clinic_053 |
| Alistipes finegoldii | 6489 | 1748 | 1448 | I_eubac_clinic_054 |
| Butyricimonas faecihominis | 17100 | 4320 | 6049 | I_eubac_clinic_054 |
| Odoribacteraceae bacterium | 4941 | 1232 | 1492 | I_eubac_clinic_054 |
| Staphylococcus caprae | 4495 | 2232 | 529 | I_eubac_clinic_055 |
| Staphylococcus lugdunensis | 11567 | 6266 | 1591 | I_eubac_clinic_055 |
| Staphylococcus caprae | 3000 | 1151 | 161 | I_eubac_clinic_056 |
| Staphylococcus lugdunensis | 16718 | 7338 | 1037 | I_eubac_clinic_056 |

| | | | | |
|---|---|---|---|---|
| Nesterenkonia flava | 12511 | 5062 | 8257 | I_eubac_clinic_057 |
| Nesterenkonia lacusekhoensis | 1245 | 460 | 689 | I_eubac_clinic_057 |
| Lactococcus lactis | 766 | 541 | 0 | I_eubac_clinic_058 |
| Staphylococcus aureus | 11461 | 7746 | 1486 | I_eubac_clinic_058 |
| Staphylococcus pasteuri | 725 | 483 | 111 | I_eubac_clinic_058 |
| Streptococcus anginosus | 21885 | 7016 | 8923 | I_eubac_clinic_059 |
| Staphylococcus caprae | 2652 | 2652 | 313 | I_eubac_clinic_060 |
| Staphylococcus caprae | 24416 | 8342 | 1277 | I_eubac_clinic_061 |
| Staphylococcus haemolyticus | 18526 | 8712 | 1731 | I_eubac_clinic_062 |
| Klebsiella variicola | 3432 | 3432 | 1705 | I_eubac_clinic_063 |
| Lactobacillus gasseri | 133 | 133 | 183 | I_eubac_clinic_063 |
| Staphylococcus aureus | 2321 | 2321 | 5286 | I_eubac_clinic_064 |
| Staphylococcus hominis | 197 | 196 | 409 | I_eubac_clinic_064 |
| Staphylococcus aureus | 20372 | 8820 | 8857 | I_eubac_clinic_065 |
| Staphylococcus aureus | 9100 | 8794 | 8799 | I_eubac_clinic_066 |
| Cloacibacterium normanense | 2493 | 1877 | 164 | I_eubac_clinic_067 |
| Corynebacterium amycolatum | 7511 | 5509 | 1077 | I_eubac_clinic_067 |
| Cutibacterium acnes | 609 | 468 | 0 | I_eubac_clinic_067 |
| Anoxybacillus kaynarcensis | 4984 | 1793 | 1431 | I_eubac_clinic_068 |
| Chitinophagaceae bacterium | 1277 | 482 | 202 | I_eubac_clinic_068 |
| Chryseobacterium moechotypicola | 3174 | 1151 | 555 | I_eubac_clinic_068 |
| Cloacibacterium normanense | 1482 | 535 | 226 | I_eubac_clinic_068 |
| Cnuibacter physcomitrellae | 1891 | 717 | 459 | I_eubac_clinic_068 |
| Corynebacterium vitaeruminis | 4646 | 1769 | 1868 | I_eubac_clinic_068 |
| Georgenia deserti | 0 | 0 | 203 | I_eubac_clinic_068 |
| Georgenia halophila | 562 | 218 | 0 | I_eubac_clinic_068 |
| Isoptericola cucumis | 444 | 164 | 186 | I_eubac_clinic_068 |
| Leuconostoc mesenteroides | 624 | 236 | 156 | I_eubac_clinic_068 |
| Linmingia china | 110 | 0 | 0 | I_eubac_clinic_068 |
| Propionibacterium westphaliense | 1579 | 949 | 1134 | I_eubac_clinic_068 |
| Pseudoclavibacter soli | 385 | 0 | 175 | I_eubac_clinic_068 |
| Sphingobacterium thermophilum | 464 | 0 | 0 | I_eubac_clinic_068 |
| Aquihabitans daechungensis | 186 | 0 | 0 | I_eubac_clinic_069 |
| Bergeyella porcorum | 503 | 154 | 0 | I_eubac_clinic_069 |
| Cloacibacterium normanense | 3547 | 1099 | 546 | I_eubac_clinic_069 |
| Cnuibacter physcomitrellae | 1514 | 481 | 476 | I_eubac_clinic_069 |
| Corynebacterium pyruviciproducens | 747 | 145 | 253 | I_eubac_clinic_069 |
| Corynebacterium ureicelerivorans | 310 | 0 | 0 | I_eubac_clinic_069 |
| Corynebacterium vitaeruminis | 3865 | 1250 | 2086 | I_eubac_clinic_069 |
| Enterococcus cecorum | 1010 | 319 | 269 | I_eubac_clinic_069 |
| Flavobacterium piscis | 233 | 0 | 0 | I_eubac_clinic_069 |
| Georgenia halophila | 263 | 0 | 0 | I_eubac_clinic_069 |
| Ilumatobacter fluminis | 112 | 0 | 0 | I_eubac_clinic_069 |
| Isoptericola cucumis | 210 | 0 | 0 | I_eubac_clinic_069 |
| Linmingia china | 107 | 0 | 0 | I_eubac_clinic_069 |
| Microbacterium sediminis | 571 | 195 | 346 | I_eubac_clinic_069 |

| | | | | |
|---|---|---|---|---|
| Microthrixaceae bacterium | 252 | 0 | 0 | I_eubac_clinic_069 |
| Moraxella osloensis | 670 | 196 | 195 | I_eubac_clinic_069 |
| Paenirhodobacter enshiensis | 315 | 0 | 0 | I_eubac_clinic_069 |
| Peribacillus butanolivorans | 0 | 0 | 110 | I_eubac_clinic_069 |
| Peribacillus loiseleuriae | 130 | 0 | 158 | I_eubac_clinic_069 |
| Peribacillus simplex | 7075 | 2226 | 1549 | I_eubac_clinic_069 |
| Staphylococcus lugdunensis | 3411 | 1127 | 926 | I_eubac_clinic_069 |
| Cutibacterium acnes | 1401 | 1279 | 151 | I_eubac_clinic_070 |
| Kocuria palustris | 7120 | 6337 | 747 | I_eubac_clinic_070 |
| Corynebacterium vitaeruminis | 1201 | 1202 | 159 | I_eubac_clinic_071 |
| Thauera aminoaromatica | 1161 | 1162 | 236 | I_eubac_clinic_071 |
| Acinetobacter bouvetii | 2112 | 1017 | 593 | I_eubac_clinic_072 |
| Acinetobacter johnsonii | 1564 | 823 | 1210 | I_eubac_clinic_072 |
| Brevibacterium mcbrellneri | 173 | 0 | 0 | I_eubac_clinic_072 |
| Brevundimonas subvibrioides | 175 | 0 | 0 | I_eubac_clinic_072 |
| Corynebacterium tuberculostearicum | 145 | 0 | 124 | I_eubac_clinic_072 |
| Cutibacterium acnes | 828 | 364 | 543 | I_eubac_clinic_072 |
| Gemella haemolysans | 853 | 400 | 368 | I_eubac_clinic_072 |
| Ligilactobacillus murinus | 794 | 386 | 284 | I_eubac_clinic_072 |
| Luteimonas aestuarii | 0 | 0 | 123 | I_eubac_clinic_072 |
| Methylobacterium adhaesivum | 0 | 0 | 283 | I_eubac_clinic_072 |
| Methylobacterium goesingense | 5174 | 2497 | 2418 | I_eubac_clinic_072 |
| Micrococcus luteus | 1323 | 639 | 542 | I_eubac_clinic_072 |
| Novosphingobium fluoreni | 384 | 160 | 107 | I_eubac_clinic_072 |
| Polaromonas jejuensis | 140 | 0 | 0 | I_eubac_clinic_072 |
| Rhodococcus corynebacterioides | 104 | 0 | 0 | I_eubac_clinic_072 |
| Sphingomonas hengshuiensis | 356 | 188 | 0 | I_eubac_clinic_072 |
| Spirosoma agri | 262 | 0 | 0 | I_eubac_clinic_072 |
| Cutibacterium acnes | 11705 | 7942 | 7092 | I_eubac_clinic_073 |
| Thiopseudomonas alkaliphila | 106 | 0 | 0 | I_eubac_clinic_073 |
| Corynebacterium tuberculostearicum | 321 | 138 | 0 | I_eubac_clinic_074 |
| Cutibacterium acnes | 342 | 190 | 0 | I_eubac_clinic_074 |
| Micrococcus lylae | 707 | 393 | 195 | I_eubac_clinic_074 |
| Pseudoglutamicibacter albus | 302 | 167 | 0 | I_eubac_clinic_074 |
| Staphylococcus aureus | 11407 | 6513 | 1686 | I_eubac_clinic_074 |
| Staphylococcus caprae | 1462 | 874 | 215 | I_eubac_clinic_074 |
| Staphylococcus haemolyticus | 762 | 420 | 105 | I_eubac_clinic_074 |
| Streptococcus humanilactis | 0 | 0 | 110 | I_eubac_clinic_075 |
| Streptococcus mitis | 0 | 0 | 322 | I_eubac_clinic_075 |
| Streptococcus oralis | 14181 | 8049 | 6839 | I_eubac_clinic_075 |
| Streptococcus toyakuensis | 0 | 0 | 239 | I_eubac_clinic_075 |
| Streptococcus chosunense | 0 | 0 | 168 | I_eubac_clinic_076 |
| Streptococcus infantis | 0 | 0 | 173 | I_eubac_clinic_076 |
| Streptococcus mitis | 0 | 0 | 180 | I_eubac_clinic_076 |
| Streptococcus oralis | 12531 | 7497 | 1484 | I_eubac_clinic_076 |
| Streptococcus toyakuensis | 0 | 0 | 175 | I_eubac_clinic_076 |

| | | | | |
|---|---|---|---|---|
| Streptococcus mitis | 0 | 0 | 160 | I_eubac_clinic_077 |
| Streptococcus oralis | 20718 | 6944 | 3564 | I_eubac_clinic_077 |
| Streptococcus toyakuensis | 0 | 0 | 123 | I_eubac_clinic_077 |
| Streptococcus pyogenes | 13491 | 7176 | 3550 | I_eubac_clinic_078 |
| Staphylococcus aureus | 7871 | 7871 | 244 | I_eubac_clinic_079 |
| Bifidobacterium dentium | 247 | 0 | 125 | I_eubac_clinic_080 |
| Capnocytophaga ochracea | 1124 | 320 | 199 | I_eubac_clinic_080 |
| Fusobacterium necrophorum | 536 | 149 | 0 | I_eubac_clinic_080 |
| Gemella morbillorum | 171 | 0 | 0 | I_eubac_clinic_080 |
| Lacticaseibacillus casei | 1356 | 382 | 328 | I_eubac_clinic_080 |
| Metamycoplasma salivarium | 330 | 0 | 0 | I_eubac_clinic_080 |
| Mogibacterium timidum | 2566 | 757 | 682 | I_eubac_clinic_080 |
| no genus brachy | 308 | 0 | 0 | I_eubac_clinic_080 |
| Olsenella phocaeensis | 168 | 0 | 0 | I_eubac_clinic_080 |
| Parvimonas micra | 7222 | 4315 | 6871 | I_eubac_clinic_080 |
| Shuttleworthia satelles | 176 | 0 | 0 | I_eubac_clinic_080 |
| Streptococcus oralis | 622 | 170 | 127 | I_eubac_clinic_080 |
| Haemophilus parainfluenzae | 9006 | 7693 | 8840 | I_eubac_clinic_081 |
| Terrahaemophilus aromaticivorans | 0 | 0 | 155 | I_eubac_clinic_081 |
| Halomonas aidingensis | 113 | 0 | 0 | I_eubac_clinic_082 |
| Streptococcus agalactiae | 12713 | 7143 | 4792 | I_eubac_clinic_082 |
| Enterobacter hormaechei | 2382 | 2383 | 169 | I_eubac_clinic_083 |
| Enterobacter asburiae | 0 | 0 | 648 | I_eubac_clinic_084 |
| Enterobacter hormaechei | 7819 | 7700 | 2985 | I_eubac_clinic_084 |
| Cutibacterium acnes | 2554 | 2554 | 654 | I_eubac_clinic_085 |
| Tepidimonas taiwanensis | 174 | 174 | 0 | I_eubac_clinic_085 |
| Corynebacterium striatum | 700 | 700 | 232 | I_eubac_clinic_086 |
| Cutibacterium acnes | 251 | 147 | 240 | I_eubac_clinic_087 |
| Fibrobacter intestinalis | 0 | 0 | 134 | I_eubac_clinic_087 |
| Hallerella succinigenes | 353 | 217 | 264 | I_eubac_clinic_087 |
| Moheibacter stercoris | 513 | 300 | 383 | I_eubac_clinic_087 |
| Staphylococcus capitis | 487 | 291 | 363 | I_eubac_clinic_087 |
| Cutibacterium acnes | 8039 | 4777 | 4182 | I_eubac_clinic_088 |
| Cutibacterium namnetense | 0 | 0 | 187 | I_eubac_clinic_088 |
| Cutibacterium acnes | 220 | 194 | 277 | I_eubac_clinic_089 |
| Staphylococcus capitis | 3408 | 1834 | 2104 | I_eubac_clinic_089 |
| Amniculibacterium aquaticum | 722 | 112 | 121 | I_eubac_clinic_090 |
| Cloacibacterium normanense | 694 | 194 | 189 | I_eubac_clinic_090 |
| Corynebacterium vitaeruminis | 730 | 232 | 279 | I_eubac_clinic_090 |
| Paenibacillus phoenicis | 1819 | 550 | 700 | I_eubac_clinic_090 |
| Pseudomonas thermotolerans | 1041 | 322 | 264 | I_eubac_clinic_090 |
| Saccharococcus thermophilus | 1324 | 383 | 284 | I_eubac_clinic_090 |
| Streptococcus australis | 0 | 0 | 119 | I_eubac_clinic_090 |
| Streptococcus infantis | 0 | 0 | 244 | I_eubac_clinic_090 |
| Streptococcus mitis | 0 | 0 | 147 | I_eubac_clinic_090 |
| Streptococcus oralis | 18707 | 5522 | 5546 | I_eubac_clinic_090 |

| | | | | |
|---|---|---|---|---|
| Streptococcus toyakuensis | 0 | 0 | 242 | l_eubac_clinic_090 |
| Pantoea agglomerans | 200 | 0 | 0 | l_eubac_clinic_091 |
| Streptococcus australis | 0 | 0 | 149 | l_eubac_clinic_091 |
| Streptococcus humanilactis | 0 | 0 | 158 | l_eubac_clinic_091 |
| Streptococcus infantis | 0 | 0 | 295 | l_eubac_clinic_091 |
| Streptococcus mitis | 0 | 0 | 201 | l_eubac_clinic_091 |
| Streptococcus oralis | 18575 | 7978 | 7629 | l_eubac_clinic_091 |
| Streptococcus pneumoniae | 0 | 0 | 114 | l_eubac_clinic_091 |
| Enterococcus cecorum | 875 | 665 | 843 | l_eubac_clinic_092 |
| Enterococcus faecium | 2174 | 1617 | 1451 | l_eubac_clinic_092 |
| Enterococcus gallinarum | 0 | 0 | 118 | l_eubac_clinic_092 |
| Enterococcus lactis | 0 | 0 | 186 | l_eubac_clinic_092 |
| Staphylococcus caprae | 1375 | 1031 | 962 | l_eubac_clinic_092 |
| Bergeyella porcorum | 566 | 115 | 115 | l_eubac_clinic_093 |
| Caldalkalibacillus uzonensis | 451 | 0 | 218 | l_eubac_clinic_093 |
| Chryseobacterium moechotypicola | 3175 | 1073 | 1031 | l_eubac_clinic_093 |
| Cloacibacterium normanense | 4091 | 1422 | 1422 | l_eubac_clinic_093 |
| Cnuibacter physcomitrellae | 1586 | 546 | 465 | l_eubac_clinic_093 |
| Corynebacterium vitaeruminis | 5937 | 2096 | 3252 | l_eubac_clinic_093 |
| Linmingia china | 1473 | 620 | 688 | l_eubac_clinic_093 |
| Microbacterium sediminis | 833 | 283 | 328 | l_eubac_clinic_093 |
| Nesterenkonia pannonica | 549 | 108 | 244 | l_eubac_clinic_093 |
| Thermoclostridium caenicola | 644 | 220 | 224 | l_eubac_clinic_093 |
| Acinetobacter baumannii | 3800 | 3801 | 5024 | l_eubac_clinic_094 |
| Acinetobacter calcoaceticus | 0 | 0 | 407 | l_eubac_clinic_094 |
| Klebsiella pneumoniae | 967 | 968 | 1993 | l_eubac_clinic_094 |
| Klebsiella quasipneumoniae | 0 | 0 | 760 | l_eubac_clinic_094 |
| Klebsiella quasivariicola | 1698 | 1698 | 398 | l_eubac_clinic_094 |
| Anoxybacillus kaynarcensis | 8721 | 2249 | 1921 | l_eubac_clinic_095 |
| Corynebacterium jeikeium | 19579 | 5695 | 6704 | l_eubac_clinic_095 |
| Meiothermus silvanus | 249 | 0 | 0 | l_eubac_clinic_095 |
| Staphylococcus haemolyticus | 703 | 165 | 120 | l_eubac_clinic_095 |
| Corynebacterium singulare | 3859 | 2167 | 1495 | l_eubac_clinic_096 |
| Corynebacterium spheniscorum | 0 | 0 | 112 | l_eubac_clinic_096 |
| Corynebacterium tuberculostearicum | 2404 | 1410 | 976 | l_eubac_clinic_096 |
| Cloacibacterium normanense | 4749 | 1362 | 1084 | l_eubac_clinic_097 |
| Cnuibacter physcomitrellae | 361 | 0 | 0 | l_eubac_clinic_097 |
| Corynebacterium vitaeruminis | 734 | 218 | 256 | l_eubac_clinic_097 |
| Dysgonamonadaceae bacterium | 1807 | 523 | 484 | l_eubac_clinic_097 |
| Georgenia deserti | 0 | 0 | 1609 | l_eubac_clinic_097 |
| Georgenia halophila | 6497 | 1093 | 435 | l_eubac_clinic_097 |
| Massiliimalia massiliensis | 3884 | 1178 | 856 | l_eubac_clinic_097 |
| Microbacterium sediminis | 2366 | 741 | 972 | l_eubac_clinic_097 |
| Prescottella equi | 3969 | 1177 | 1084 | l_eubac_clinic_097 |
| Rhodobacter flagellatus | 784 | 234 | 120 | l_eubac_clinic_097 |
| Staphylococcus caprae | 2055 | 617 | 617 | l_eubac_clinic_097 |

| | | | | |
|---|---|---|---|---|
| Staphylococcus aureus | 16433 | 2813 | 8711 | I_eubac_clinic_098 |
| Streptococcus pneumoniae | 2297 | 487 | 0 | I_eubac_clinic_099 |
| Streptococcus pseudopneumoniae | 35386 | 7215 | 8682 | I_eubac_clinic_099 |
| Cutibacterium acnes | 137 | 0 | 0 | I_eubac_clinic_100 |
| Neisseriaceae bacterium | 356 | 0 | 0 | I_eubac_clinic_100 |
| Staphylococcus capitis | 35907 | 8710 | 8479 | I_eubac_clinic_100 |
| Staphylococcus caprae | 0 | 0 | 224 | I_eubac_clinic_100 |
| Staphylococcus aureus | 41395 | 8618 | 8768 | I_eubac_clinic_101 |
| Staphylococcus aureus | 0 | 0 | 138 | NegK-20231103 |
| Anoxybacillus kaynarcensis | 0 | 0 | 496 | NegK-240404 |
| Caldibacillus hisashii | 0 | 0 | 256 | NegK-240404 |
| Cnuibacter physcomitrellae | 536 | 279 | 0 | NegK-240404 |
| Corynebacterium vitaeruminis | 3626 | 2014 | 0 | NegK-240404 |
| Cutibacterium acnes | 2302 | 1265 | 0 | NegK-240404 |
| Enterococcus cecorum | 0 | 0 | 323 | NegK-240404 |
| Isoptericola cucumis | 238 | 0 | 0 | NegK-240404 |
| Lawsonella clevelandensis | 548 | 309 | 0 | NegK-240404 |
| Lihuaxuella thermophila | 0 | 0 | 860 | NegK-240404 |
| Linmingia china | 642 | 343 | 0 | NegK-240404 |
| Sphingorhabdus rigui | 1522 | 846 | 0 | NegK-240404 |
| Staphylococcus caprae | 422 | 245 | 0 | NegK-240404 |
| Staphylococcus warneri | 5133 | 2726 | 0 | NegK-240404 |
| Thermicanus aegyptius | 0 | 0 | 950 | NegK-240404 |
| Actinomycetales bacterium | 110 | 110 | 233 | NegK-Extrakt-240705 |
| Bifidobacterium santillanense | 0 | 0 | 367 | NegK-Extrakt-240705 |
| Moraxella osloensis | 145 | 145 | 160 | NegK-Extrakt-240705 |

**Supplementary Table 5.** Cohen's *Kappa* calculations for different raters.

| rater: Illumina+clinician review | rater: ONT+clinician review | | |
|---|---|---|---|
| *categories* | found only the likely pathogen | found likely pathogen and contamination | found only contamination |
| found only the likely pathogen | 44 | 5 | 0 |
| found likely pathogen and contamination | 12 | 16 | 0 |
| found only contamination | 0 | 0 | 24 |

**Weighted Kappa =** 0.80517
**SE of weighted kappa =** 0.0465

| rater: Illumina subsample+clinician review | rater: ONT+clinician review | | |
|---|---|---|---|
| *categories* | found only the likely pathogen | found likely pathogen and contamination | found only contamination |
| found only the likely pathogen | 52 | 5 | 0 |
| found likely pathogen and contamination | 4 | 16 | 0 |
| found only contamination | 0 | 0 | 24 |

**Weighted Kappa =** 0.89582
**SE of weighted kappa =** 0.03462
